# Problematic usage of the internet and eating disorders: a multifaceted, systematic review and meta-analysis

**DOI:** 10.1101/2020.08.20.20177535

**Authors:** Konstantinos Ioannidis, Charlotte Taylor, Leah Holt, Kate Brown, Christine Lochner, Naomi A Fineberg, Ornella Corazza, Samuel R Chamberlain, Andres Roman-Urrestarazu, Katarzyna Czabanowska

## Abstract

Eating disorders are widespread illnesses with significant impact. There is growing concern about how those at risk of eating disorders overuse online resources to their detriment. We conducted a pre-registered systematic review and meta-analysis of studies examining Problematic Usage of the Internet (PUI) and eating disorders. The meta-analysis comprised n = 32,295 participants, in which PUI was correlated with significant eating disorder psychopathology Pearson r = 0.22 (s.e. = 0.04, p< 0.001), body dissatisfaction r = 0.16 (s.e. = 0.02, p< 0.001), drive-for-thinness r = 0.16 (s.e. = 0.04, p< 0.001) and dietary restraint r = 0.18 (s.e. = 0.03). Effects were not moderated by gender, PUI facet or study quality. Results are in support of PUI impacting significantly on vulnerable populations towards the development or maintenance of eating disorder psychopathology; males may be equally vulnerable to these potential effects. Prospective and experimental studies in the field suggest that small but significant effects exist and may have accumulative influence over time and across all age groups. Those findings are important to expand our understanding of PUI as a multifaceted concept and its impact on multiple levels of ascertainment of eating disorder psychopathology.

## 1. Introduction

From the inception of the world-wide-web in the 1980s, to the global domination of multifaceted online-based applications of today, the Internet has shaped our lives irreversibly (Fineberg et al., 2018; Young, 1998). Problematic usage of the internet (PUI) is an umbrella term referring to maladaptive engagement in online activities known to be associated with marked functional impairment (Fineberg et al., 2018). PUI describes engagement with a variety of online activities, including: gaming, use of social networking sites (SNS) and streaming platforms, online gambling, compulsive online buying, online pornography and cyberbullying victimization (Fineberg et al., 2018; Ioannidis et al., 2018). The online activities associated with PUI are excessive and functionally impairing, e.g. leading to poorer health, social, vocational or academic outcomes or lower quality of life (Fineberg et al., 2018; Ioannidis et al., 2019a; Kuss et al., 2014). Other less examined areas of online engagement include the excessive use of dating, weight control and fitness applications (Apps), and the excessive consumption of health resources to alleviate health anxiety. In the last decade, the number of studies reporting PUI outcomes on eating disorder and related psychopathology and has increased exponentially (Figure 1).

**Figure 1.**
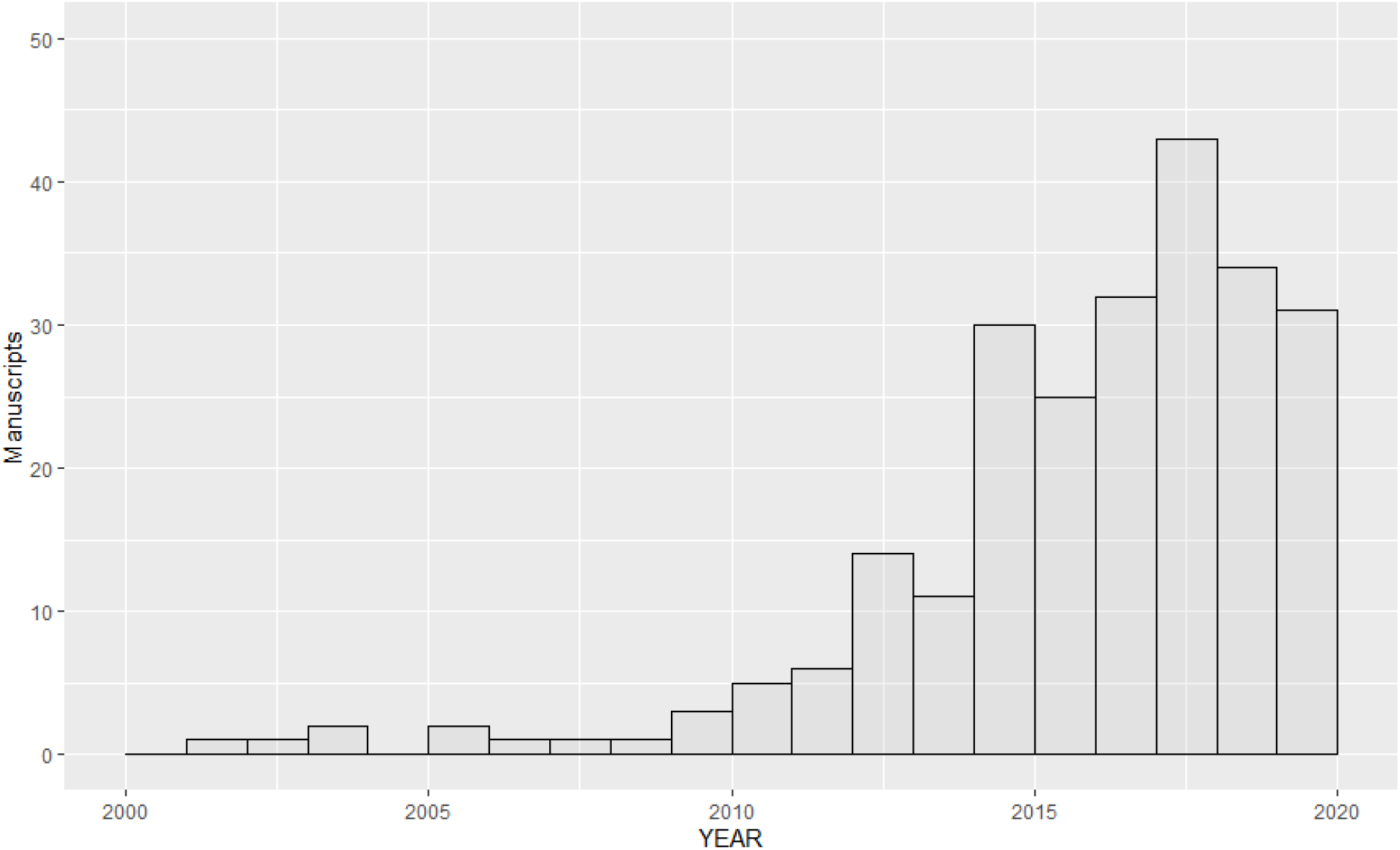
Histogram representing the number of studies examining the problematic use of internet and eating disorders. X-axis: period covered; to note, the 2020 bar covers a period only Jan-May 2020. Y-axis, number of manuscripts identified as relevant titles through Pubmed search.

### 1.1 Problematic usage of the internet and eating disorders

The influence of media on eating disorders (EDs) has been known for many years (Martínez-González et al., 2003). In a sample of 77 studies, general mass media exposure negatively impacted women’s body dissatisfaction (d = –0.28), internalization of the thin ideal (d = –.39) and eating behaviors and beliefs (d = –0.30) (Grabe et al., 2008). Recently, there is growing concern about how new technology, including but not exclusively SNS use, may be affecting the development or perseverance of EDs (Mingoia et al., 2017; Slater et al., 2019; Tiggemann and Slater, 2013; Wilksch et al., 2020). Indeed, online media may have a stronger effect on self-objectification parameters as compared to traditional media (e.g. TV) (Karsay et al., 2018).

Various SNS have increasingly been used as sources of information and reinforcement regarding beauty and fitness ideals. On these platforms, users may be inclined to compare their physical attributes to unrealistic and unattainable body ideals (Tiggemann and Zaccardo, 2015; Wick and Keel, 2020). Concerns have been raised on how the use of SNS may be affecting the development or perseverance of eating disorders (Mingoia et al., 2017; Slater et al., 2019, 2017; Smith et al., 2013; Tiggemann and Miller, 2010; Tiggemann and Slater, 2017, 2013). Recent SNS trends, such as “fitspiration”, a word arising from the amalgamation of the words “fitness” and “inspiration”, strongly idealise very thin and much toned body images, whereas other SNS “thinspiration” or “bonespiration” content underline physical changes in response to specific eating disordered behaviours such as restrictive eating and extreme weight loss (Carrotte et al., 2015; Giorgetti et al., 2020; Talbot et al., 2017). Indeed, SNS use was found to prospectively predict an increase in drive for thinness two years later (Tiggemann and Slater, 2017), supporting a hypothetical causal role for engaging with social media on ED cognitions. Specific populations studied for SNS overuse have been found to struggle on an executive and inhibitory control level (impulse control), in similarity to those suffering from addictive behaviors (Wegmann et al., 2020, 2018b, 2018a). Indeed, Social-network-use disorder has been considered for inclusion in the new version of the International Classification of Diseases (ICD-11) as an ‘other specified disorders due to addictive behaviors’ (Brand et al., 2020). The relationship between online behaviors and impulsive ED behaviors (e.g. bingeing, purging) has also been demonstrated (Butkowski et al., 2019; Holland and Tiggemann, 2017; Melioli et al., 2015; Smith et al., 2013; Tao, 2013; Tao and Liu, 2009; Wilksch et al., 2020)).

A number of studies have used an ‘Internet Addiction’ or ‘PUI’ paradigm and associated ‘global’ problematic usage of the internet, with eating disorder psychopathology, (Alpaslan et al., 2015; Canan et al., 2014; Çelik et al., 2015; Fernández-villa et al., 2015; Martínez-González et al., 2014; Rodgers et al., 2013; Tao, 2013). Other studies examined other specific facets of usage, for example use of calorie tracking Apps (Embacher Martin et al., 2018; Simpson and Mazzeo, 2017), dating Apps (Griffiths et al., 2018b; Rodgers et al., 2019; Tran et al., 2019), cyberbullying victimization (Kelly et al., 2018; Kenny et al., 2018; Marco et al., 2018; Olenik-Shemesh and Heiman, 2017; Pistella et al., 2019), and consumption of eating disorder promoting content (pro-ED content, within SNS or in the global online environment), like ‘fitspiration’, ‘thinspiration’ (Griffiths et al., 2018a) or pro-ED (pro-eating-disorder content) (Rodgers et al., 2012). In reality, many of those facets of usage overlap and co-occur with each other while we interact with the online milieu (Figure 2). For example, someone may be posting calorie counting ‘achievements’ using a smartphone App, while consuming pro-ED content using SNS and experiencing cyberbullying victimization through interactions with fellow users. Those different facets of PUI may or may not have differential impact on precursors, risk factors and eating disorder clinical outcomes; no previous study has synthesized available knowledge across the various facets to investigate those associations.

**Figure 2.**
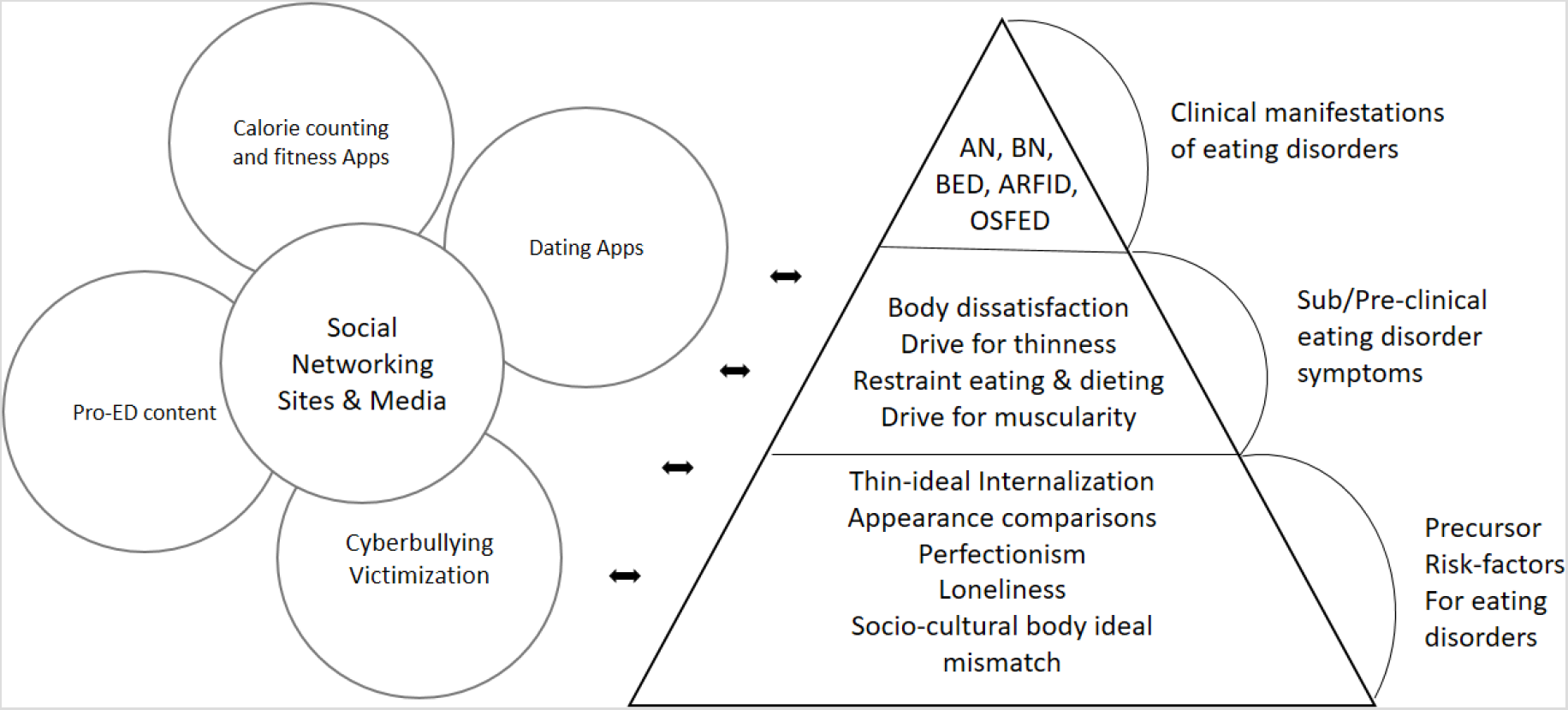
Schematic hypothesized representation of overlapping online activities and their potential relationship with precursors, pre-clinical and clinical manifestations of eating disorders.

Previous meta-analysis has examined the relationship between use of SNS and the internalization of the thin ideal (ITI) (Mingoia et al., 2017). They assessed six studies examining this relationship and found SNS significantly associated with ITI with r = 0.18 (s.e. 0.12–0.23). The study also compared outcomes measuring broad use of SNSs and outcomes measuring SNS use solely as a function of specific appearance-related features (e.g., posting or viewing photographs). The use of appearance-related features had a stronger relationship with the ITI than broad use of SNSs. The ITI is a process which involves the development of a positive association and perceived reward with an unattainable model of thinness, often associated with attractiveness and success (Cash and Pruzinsky, 2004). It is considered a risk factor for the development of eating disorders, together with restrained eating (Thompson and Stice, 2001). While historically mass media have been the culprits of incomplete, inaccurate and biased content, modern online media, have taken this to the extreme, with dozens of applications easily accessible to publicize and consume image-oriented, biased and manipulated content (Lonergan et al., 2019).

### 1.2 Aims and hypotheses

In this study we aim to provide a quantitative analysis of all available evidence linking PUI and eating disorder and related psychopathology. In doing so, our review aims to answer how PUI influences eating disorder and related psychopathology, and what the moderating parameters influencing this relationship are.

## 2. Methods

Our systematic review and meta-analysis protocol was pre-registered electronically [initially on the 6^th^ of March 2020 and updated on the 3^rd^ August 2020 to include details of our meta-analysis strategy] and published online on the PROSPERO International prospective register of systematic reviews and followed the PRISMA-P recommendations for systematic reviews and meta-analysis (Shamseer et al., 2015) [Available from: https://www.crd.york.ac.uk/prospero/display_record.php?ID=CRD42020172410].

### 2.1 Search strategy

Our search and screening strategy is outlined in Figure 3.

**Figure 3.**
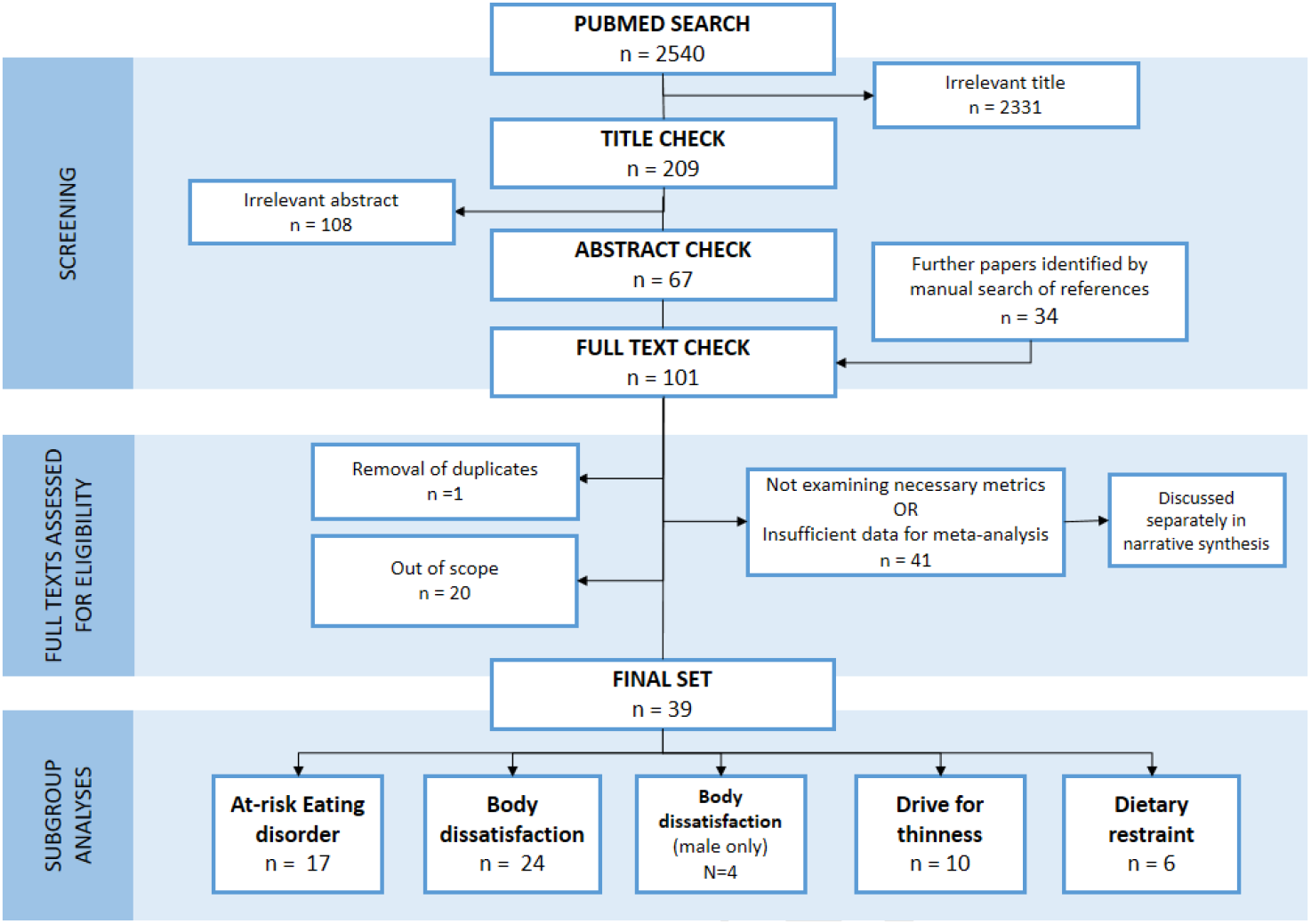
PRISMA flowchart; search results, screening and study numbers entering meta-analysis of problematic usage of the internet and eating disorders

The search string was determined by consensus amongst the co-authors. A Pubmed search was conducted with the following string: (((internet addiction[Title/Abstract] OR problematic internet*[Title/Abstract] OR internet[Title/Abstract] OR online[Title/Abstract] OR gaming[Title/Abstract] OR videogaming[Title/Abstract] OR video-gaming[Title/Abstract] OR social media[Title/Abstract] OR cyberbullying [Title/Abstract] OR cyber-bullying*[Title/Abstract] OR cyberharassment[Title/Abstract] OR cyber-harassment[Title/Abstract] OR smartphone*[Title/Abstract] OR phone*[Title/Abstract])) AND (eating disorder*[Title/Abstract] OR body dissatisf*[Title/Abstract] OR disordered eat*[Title/Abstract] OR weight[Title/Abstract] OR BMI[Title/Abstract] OR BDD [Title/Abstract] OR body dysm*[Title/Abstract])) AND (exercise OR physical activi* OR fitness* OR thinspiration* OR sport OR gym OR body image OR disordered eat* OR body shaming*)]. We included Body-Dysmorphic Disorder (BDD) in our inclusion strategy, as individuals with BDD share body dissatisfaction symptoms with those who have eating disorders (Hrabosky et al., 2009).

### 2.2 Inclusion criteria

We included all studies that (a) were published in scholarly peer reviewed journals between 1993 and May 2020; (b) were written in English or Spanish or provided an English translation; (c) examined participants with PUI (used in its wider meaning to include the full spectrum of ‘addictive use of the internet’, ‘problematic internet use’, ‘overuse of social media’, ‘cyberbullying victimization’, and ‘internet gaming disorder’) and (d) included measures of eating disorders, disordered eating, or body dissatisfaction or measures of excessive exercise.

### 2.3 Exclusion criteria

We excluded all studies that a) did not provide quantitative analysis results and b) were published only in the grey literature (including conference papers, non-peer reviewed publications, doctoral theses; as these sources are not necessarily subject to the same journal-level rigorous peer-review procedures as non-grey literature).

### 2.4 Data collection and analysis

Data were extracted from the original papers or were provided by the authors of each study. Information from the included studies, where available, was recorded and different types of data were extracted from each study including (a) a geographical determinant in which the data collection occurred; (b) key demographics of the participants (age as categorized by mean age reported in the sample, also grouped as ‘Children and Adolescents (< 18yrs)’, ‘Youth (18–25yrs)’, ‘Adults (25+yrs)’); a percentage/ratio distribution of gender; (c) body mass index (BMI) mean and SD for the whole sample; (d) education descriptors of the sample; (e) operationalization of PUI including the assessment instrument that was used and the clinical significance cut-off; (f) operationalization of eating disorder symptomatology including the instruments used and significance cut-offs; (g) reported psychiatric co-morbidities in the sample; (h) Pearson correlation coefficients between PUI and eating disorder measures; (i) measures of body dysmorphic disorder, (j) measures of exercise addiction, and (k) study quality scores. The quality assurance control was performed independently using the Effective Public Health Practice Project (EPHPP) (Armijo-Olivo et al., 2012), a standardized instrument for the assessment of study quality in quantitative research, and by an ad-hoc instrument, specifically designed for the purposes of this study. All studies that entered the meta-analysis received a rating of ‘Strong’, ‘Moderate’ or ‘Weak’ from the EPHPP assessment by two raters. One-way Intra-class correlation between raters was excellent 0.88 (0.802–0.936) for the EPHPP. All papers in scope were assessed against the ad-hoc instrument and received a score ranging between 0 and 22, with higher scores indicating higher quality (see Supplement TABLE S1). Details of the ad-hoc instrument are also presented (see Supplement TABLE S2). The full list and references of studies that entered the systematic review are reported in supplemental TABLE S1. The metrics included in the meta-analysis are presented in detail in the supplemental file.

### 2.5 Analysis methodology

#### 2.5.1 Meta-analysis strategy

In our pre-registered analysis plan, we proposed ascertaining eating disorder psychopathology as a group level difference between exposed group (PUI in various facets) versus controls. We anticipated a high heterogeneity of outcome measures that would require a post-hoc judgment about whether a quantitative synthesis is possible. We aimed to produce quantitative statistics in respect to clinical parameters (e.g. epidemiological data of prevalence/incidence for the disorders in scope) and other metrics if available (e.g. reported degree of comorbidities, BMI, activity levels, quality of life), however those data proved to be scarce in the papers in scope. Instead, we identified substantial data to quantitatively describe various aspects of eating disorder psychopathology in observational studies using correlation statistics, between PUI-facet-specific exposure (e.g. Problematic usage of the internet, or Gaming disorder sufferers or SNS use or Cyberbullying victimization) and eating disorder symptomatology.

#### 2.5.2 Metrics used for the construction of a uniform and comparable effect

The vast majority of studies reviewed here reported correlation statistics using Pearson’s r correlation coefficient. We extracted Pearson’s r correlation statistics from cross-sectional observational studies that examined the relationship between PUI facet and eating disorder metrics. We excluded studies in which we did not have variables comparable with Pearson’s r. Authors were contacted to provide information on their studies and given the opportunity to provide raw data and those studies were also included in the meta-analysis. Studies without available and comparable metrics are discussed separately, but not entered in meta-analysis. We, therefore, excluded studies that reported logistic regression coefficients, partial correlation statistics, pathway coefficients as those coefficients are either non-compatible with our approach, or would require a substantial degree of data transformation under assumptions with significant degree of uncertainty. However, if those studies also reported Pearson’s r they were included. Finally, we excluded studies that reported prospective or cross-lagged correlations, not because we did not consider them extremely important to understanding the associations under scope, but rather because they were incompatible with our meta-analytical approach.

#### 2.5.3 Internet exposure metrics under consideration

While there is substantial variability in the internet exposure metrics, we focused on those that are aimed to ascertain a general engagement on the online medium. Exposure metrics were considered equally, despite arguable differences in quality between them. For studies that included an established PUI measure we utilized the total score as exposure measure. In those studies that reported multiple metrics of engagement with online media, we chose the most general metric (e.g. time spend on the medium) that to our view best ascertains the overarching exposure to the online environment and is relatively ‘neutral’ in relation to eating disorder specific behaviors. In previous works, those ED-influenced metrics have been shown to formulate stronger correlations with ED psychopathology (Mingoia et al., 2017) and therefore meta-analysing those associations to explore the relationship between online behaviors and eating disorder symptoms may inadvertently fail to circular reasoning. For that reason, we specifically tried to avoid utilizing metrics that did not appear to be ‘ED neutral’ e.g. ‘how much time do I spend comparing my body-shape to others on Instagram’, as those metrics were more likely to represent an online manifestation of ED psychopathology, rather than a measure of exposure to the online environment. More details about internet use metrics included in the meta-analysis is presented in the supplemental file (∫ S1).

#### 2.5.4 Eating disorder symptoms metrics under consideration

While there is substantial variability in the ED metrics reported as outcomes of the studies in scope, we focused on specific domains that provided enough data for a quantitative approach and assessed relatively uniform dimensions of symptoms and behavior:

1. At-risk-eating disorders: we used total scores of Eating Disorder Examination Questionnaire (EDE-Q) (Fairburn and Beglin, 1994), Eating Disorder Inventory (EDI) (Garner et al., 1983), Eating Attitudes Test (EAT-26 or EAT-40) (Garner et al., 1982), and the SCOFF Eating Disorder Questionnaire (Luck et al., 2002), as those instruments aimed to ascertain a global degree of ED psychopathology. While specific cut-offs can be used to ascertain clinical significance with a margin of certainty, due to the fact that our approach aims to establish correlations within all degrees of severity, we consider this analysis as providing a compound metric of ‘at-risk eating disorders’.
2. Body dissatisfaction (BD) is a transdiagnostic symptom, core to eating disorders and BDD: we used the EDI body dissatisfaction subscale (EDI-BD) score, Body Image Avoidance Questionnaire (BIAQ) score (Rosen et al., 1991), Body Shape Questionnaire (BSQ) score (Cooper et al., 1987), EDE-Q Shape concern subscale, the Contour Drawing Rating Scale (CDRS) difference score (Thompson and Gray, 1995), the Body Attitudes Test (BAT) body dissatisfaction subscale (BAT-BD) (Probst et al., 1995), the Body Image States Scale (BISS) (Cash et al., 2002), and the New Zealand Attitudes and Values Study (NZAVS) questionnaire battery body dissatisfaction scale (NZAVS-BD) (Stronge et al., 2015).
3. Drive-for-Thinness (DT) is a transdiagnostic ED cognition, predominantly present in anorexia nervosa and bulimia nervosa: we used the EDI drive-for-thinness subscale (EDI-DT) and the EDE-Q weight concern subscale.
4. Dietary restraint (DR) is a transdiagnostic ED symptom, predominantly present in anorexia nervosa and bulimia nervosa: we used the Dutch Eating Behavior Questionnaire (DEBQ) dietary restraint subscale (DEBQ-R) (Strien et al., 1986) and the EDE-Q restraint subscale.

More details about the ED metrics included in the meta-analysis are presented in the supplemental file (∫ S2.1–2.6).

#### 2.5.5 Effect sizes pre-processing

Effect sizes of individual studies were calculated using the Fisher’s r-to-z transformation (Hedges and Vevea, 1998), in which a summary Fisher’s z is calculated and entered into the meta-analysis to calculate a summary effect, before being converted back to r and presented as a weighted average of these transformed scores. A positive effect size indicates a higher level of ED pathology is associated with an increase in the specific PUI facet exposure. Correlation values of 0.10, 0.30, and 0.50 represent small, moderate, and large effects, respectively (Cohen, 1988). We used Random-effects as appropriate for this methodological approach, pooling effects from different populations (Hedges and Vevea, 1998; Quintana, 2015).

#### 2.5.6 Data-analysis

Meta-analysis was conducted where at least four datasets were available for a given type of ED domain. For convenience, all meta-analysis plots were shown such that positive values on the X-axes indicated a higher degree of ED psychopathology. We first conducted an exploratory analysis of influence to identify outliers. During this process, we excluded studies that were identified as highly influential (Cook’s d influence > 2 SD above domain mean) and were of weak quality score (Ioannidis et al., 2019b). Data were analyzed using statistical software R version 3.5.3. Meta-analysis was performed using packages of “robumeta” and “metafor” (Fisher et al., 2017; Viechtbauer, 2020). The R code used for this analysis is shared in the supplement to support reproducible research. Moderator analysis (all models were meta-regression) was conducted for age, gender, geographical region of reporting, and quality scores.

## 3 Results

The initial search yielded 2,540 results. Those provided 209 relevant titles for further review. The majority of these were excluded on abstract review, due to being out of scope (e.g. papers not measuring ED symptoms, or unrelated to PUI). This yielded 101 papers possibly eligible for inclusion including manual identification through references checks for further papers within scope. After excluding papers that were out-of-scope from going through full texts a final set of 80 studies, including a total of 145,809 participants from 21 countries was identified, 39 of which were eligible for meta-analysis (Figure 4).

**Figure 4.**
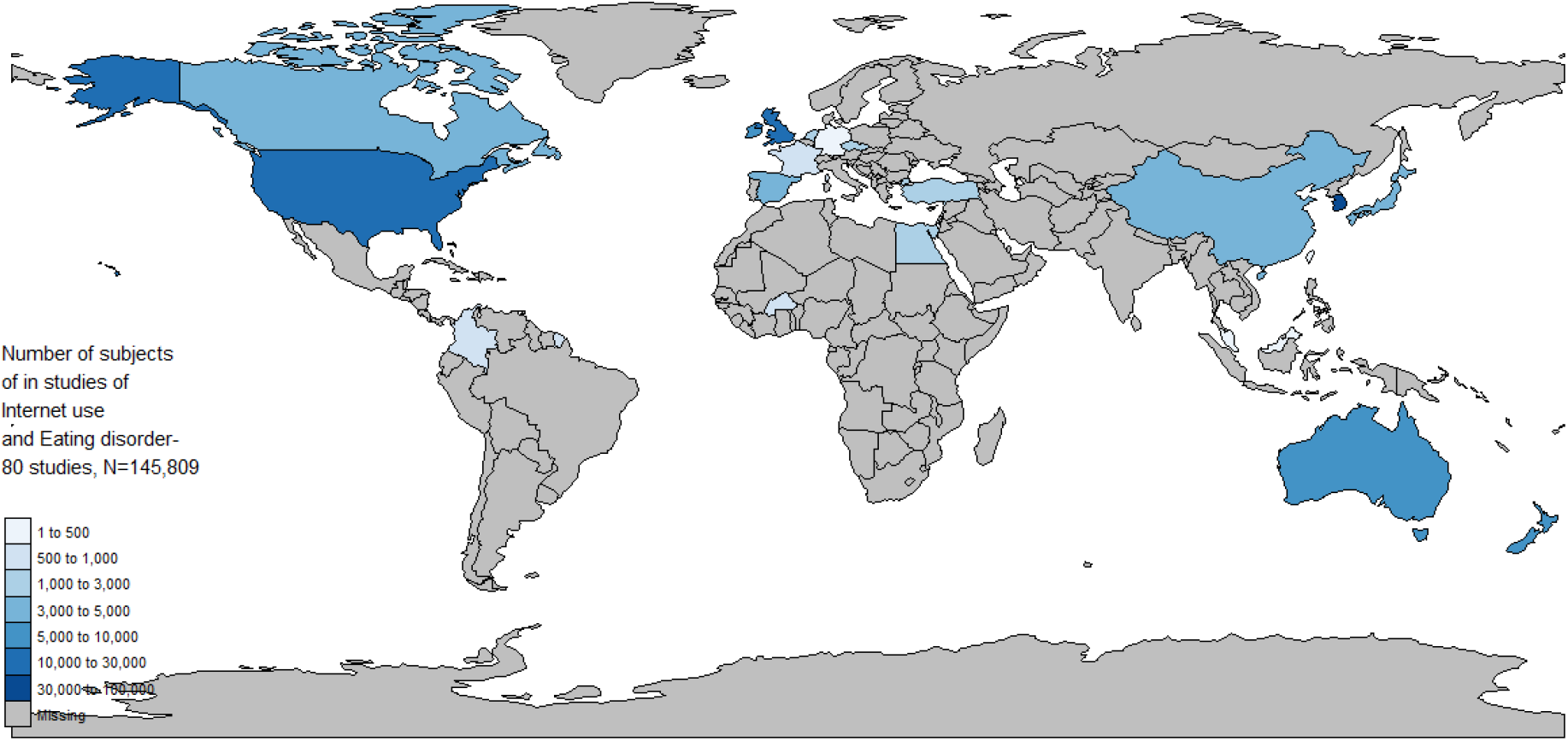
Global choropleth map indicating the number of study participants in different countries; studies included here are only those as screened eligible for the systematic review.

Out of those, 39 studies entered the meta-analysis, with a total 32,295 number of participants.

### 3.1 Heterogeneity measures

Heterogeneity measures of tau-squared (estimated amount of total heterogeneity), I^2^ (total heterogeneity / total variability) and Q-test p-values are reported separately for each sub-analysis in TABLE 1.

**Table 1.**
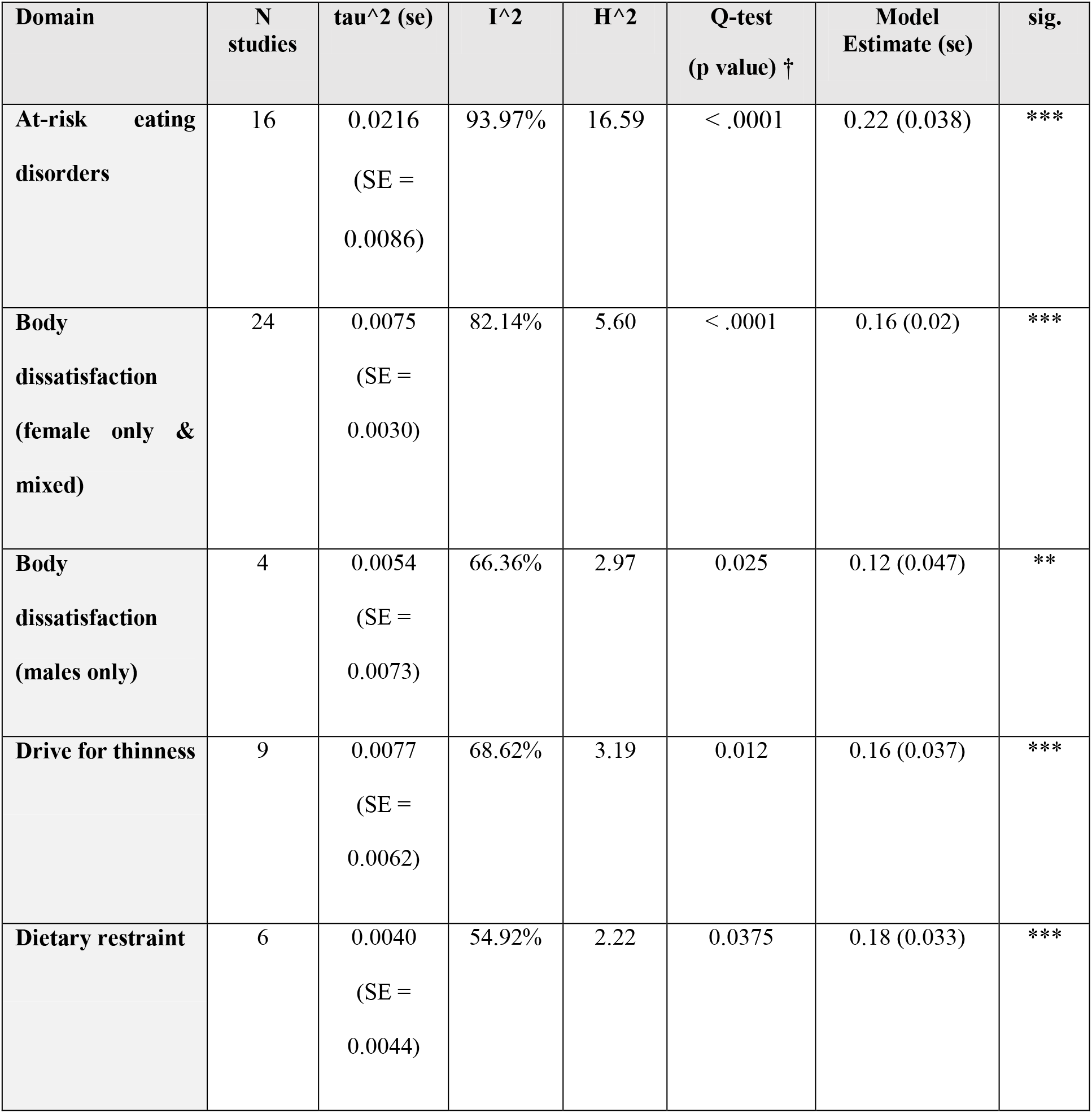
Heterogeneity and model estimate measures for different cognitive domains. ***tau^2***: estimated amount of total heterogeneity; ***I^2***: (total heterogeneity / total variability); ***H^2***: (total variability / sampling variability); ***Q-test***: Test for Heterogeneity; meta-analysis was done using random-effects model using REML. ***REML***: Restricted maximum-likelihood estimator

| Domain | N studies | $\tau^2$ (se) | $I^2$ | $H^2$ | Q-test (p value) † | Model Estimate (se) | sig. |
| --- | --- | --- | --- | --- | --- | --- | --- |
| At-risk eating disorders | 16 | 0.0216<br>(SE = 0.0086) | 93.97% | 16.59 | < .0001 | 0.22 (0.038) | *** |
| Body dissatisfaction (female only & mixed) | 24 | 0.0075<br>(SE = 0.0030) | 82.14% | 5.60 | < .0001 | 0.16 (0.02) | *** |
| Body dissatisfaction (males only) | 4 | 0.0054<br>(SE = 0.0073) | 66.36% | 2.97 | 0.025 | 0.12 (0.047) | ** |
| Drive for thinness | 9 | 0.0077<br>(SE = 0.0062) | 68.62% | 3.19 | 0.012 | 0.16 (0.037) | *** |
| Dietary restraint | 6 | 0.0040<br>(SE = 0.0044) | 54.92% | 2.22 | 0.0375 | 0.18 (0.033) | *** |

### 3.2 At risk eating disorders

In performing meta-analysis for the at-risk eating disorders domain we removed one of the studies that was highly influential (reporting an extremely positive Pearson correlation r = 0.77, Cook’s d > 2 S.D. above mean of the cohort of studies, n = 314) and of weak quality (Çelik et al., 2015). By doing so, the pooled estimate result is reported more conservatively. In the final sample. sixteen studies were included (n = 13168 study participants) (Alpaslan et al., 2015; Bair et al., 2012; Canan et al., 2014; Cañon Buitrago et al., 2016; Cohen and Blaszczynski, 2015; Eckler et al., 2017; Fernández-villa et al., 2015; Griffiths et al., 2018a; Kamal and Kamal, 2018; Mabe et al., 2014; Marco et al., 2018; Quesnel et al., 2018; Rodgers et al., 2013; Simpson and Mazzeo, 2017; Walker et al., 2015; Zeeni et al., 2018) with a small pooled estimate effect Pearson’s r = 0.22 (s.e. = 0.037, p< 0.001) (Figure 5).

**Figure 5.**
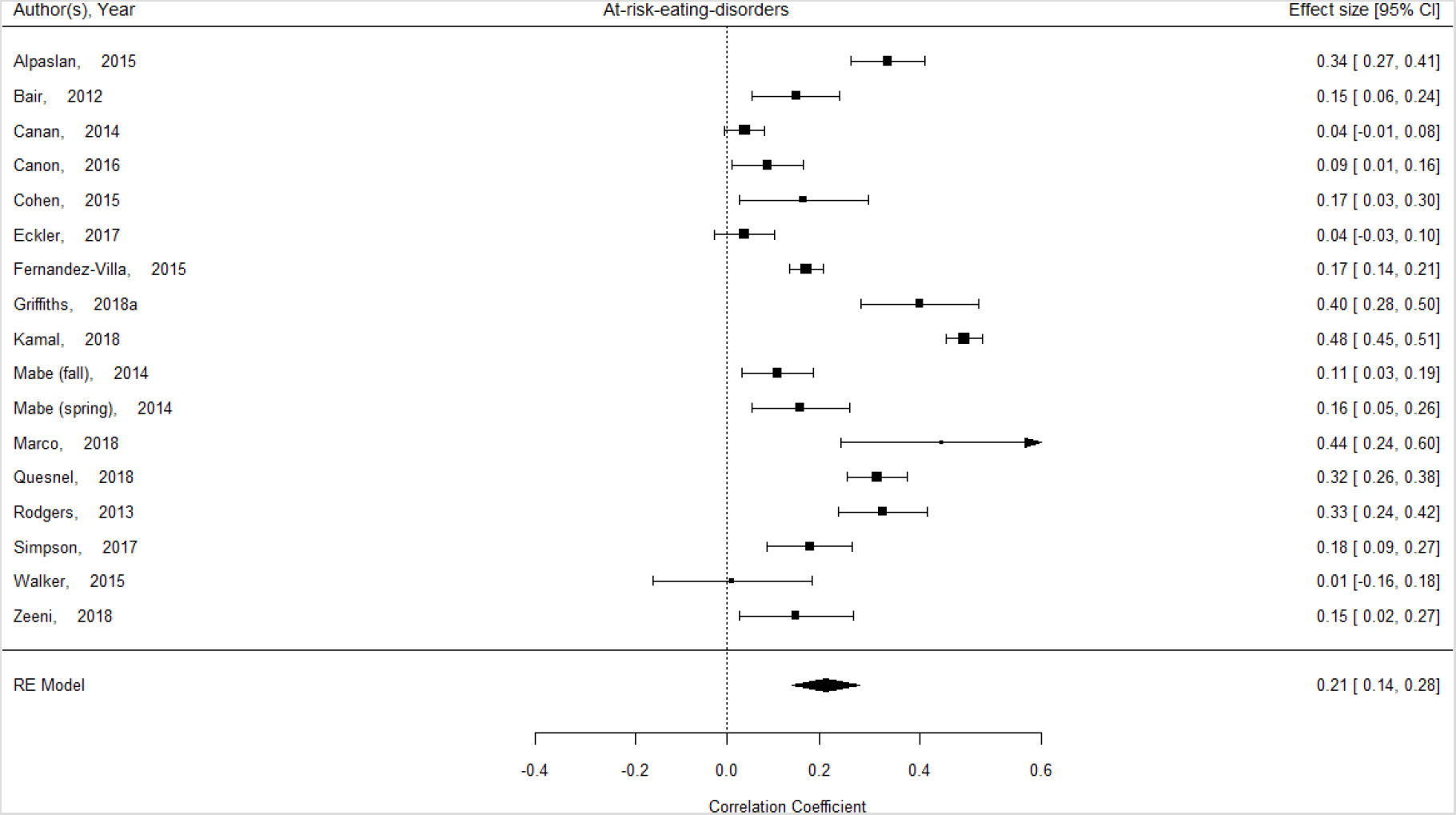
Meta-analysis of Problematic usage of the internet (PUI) by ‘at-risk eating disorders’, using Pearson correlation. Positive Pearson correlation indicates that a higher score of PUI is associated with higher risk for eating disorders. Correlation values of 0.10, 0.30, and 0.50 represent small, moderate, and large effects, respectively

### 3.3 Body dissatisfaction

In performing meta-analysis for the body dissatisfaction domain, we identified 24 studies in scope, four of which reported data separately for males and females. Pooled estimates are reported together (all studies; n = 15494) (Bair et al., 2012; Butkowski et al., 2019; Cohen and Blaszczynski, 2015; de Vries et al., 2019, 2016; Eckler et al., 2017; Hendrickse et al., 2017; Hummel and Smith, 2015; Levinson et al., 2017; McLean et al., 2015; Olenik-Shemesh and Heiman, 2017; Rodgers et al., 2020, 2019, 2013; Simpson and Mazzeo, 2017; Smith et al., 2013; Stronge et al., 2015; Tao, 2013; Terhoeven et al., 2020; Tiggemann and Miller, 2010; Veldhuis et al., 2018; Verbist and Condon, 2019; Xiaojing, 2017; Zeeni et al., 2018) and for males separately (Rodgers et al., 2020, 2019; Stronge et al., 2015; Xiaojing, 2017). The pooled estimate from all 24 studies was small, r = 0.16 (s.e. = 0.02, p< 0.001). A secondary sub-analysis of female only studies (14 studies had only female samples or reported females separately) had a similar pooled estimate, r = 0.14 (s.e. = 0.03) (Figure 6).

**Figure 6.**
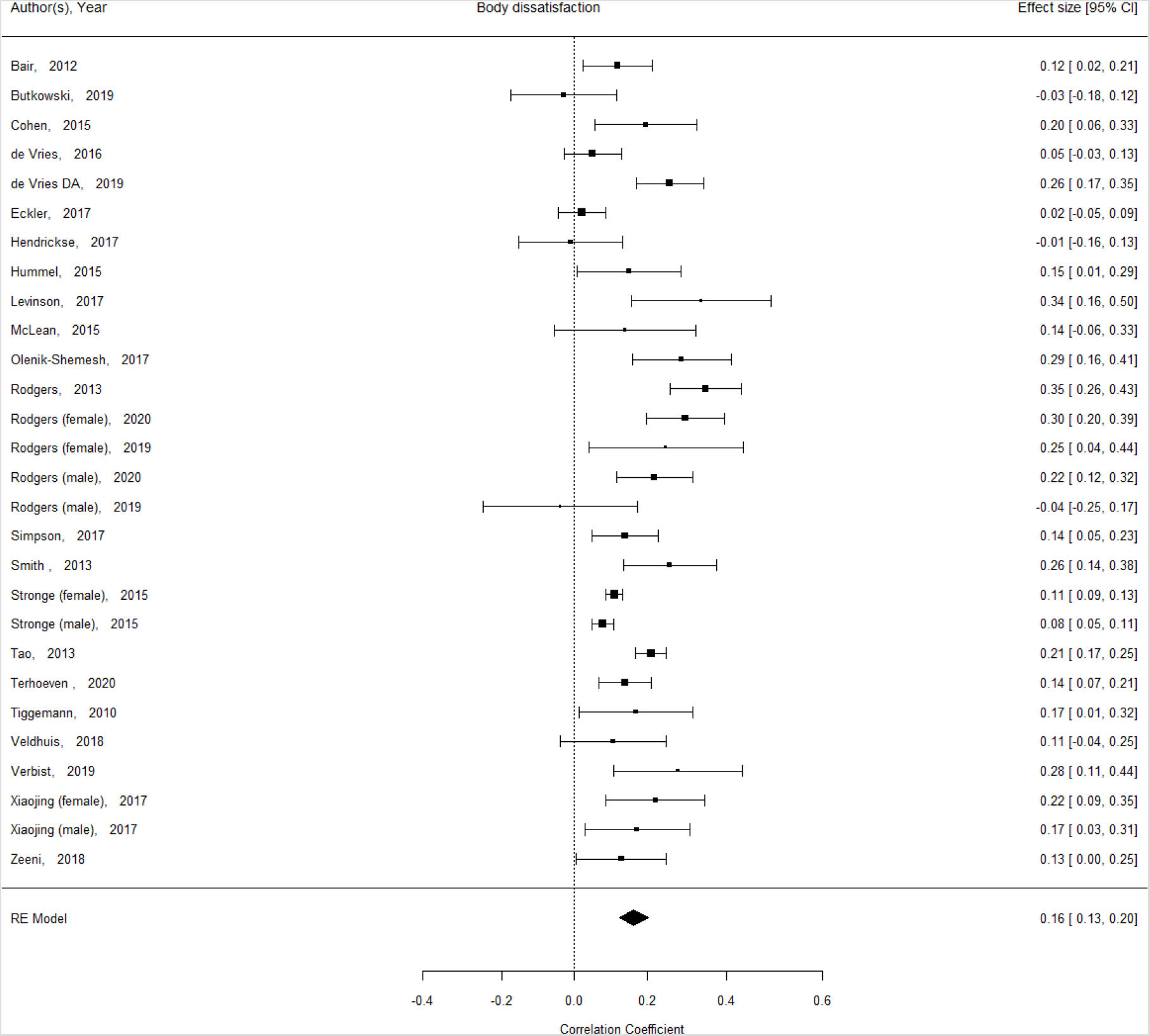
Meta-analysis of Problematic usage of the internet (PUI) by ‘body dissatisfaction’, using Pearson correlation. Positive Pearson correlation indicates that a higher score of PUI is associated with higher body dissatisfaction. Correlation values of 0.10, 0.30, and 0.50 represent small, moderate, and large effects, respectively

Four studies included separate data for n = 4749 males (Rodgers et al., 2020, 2019; Stronge et al., 2015; Xiaojing, 2017) which provided a small pooled estimate of r = 0.12 (s.e. = 0.047, p< 0.01) (Figure 7).

**Figure 7.**
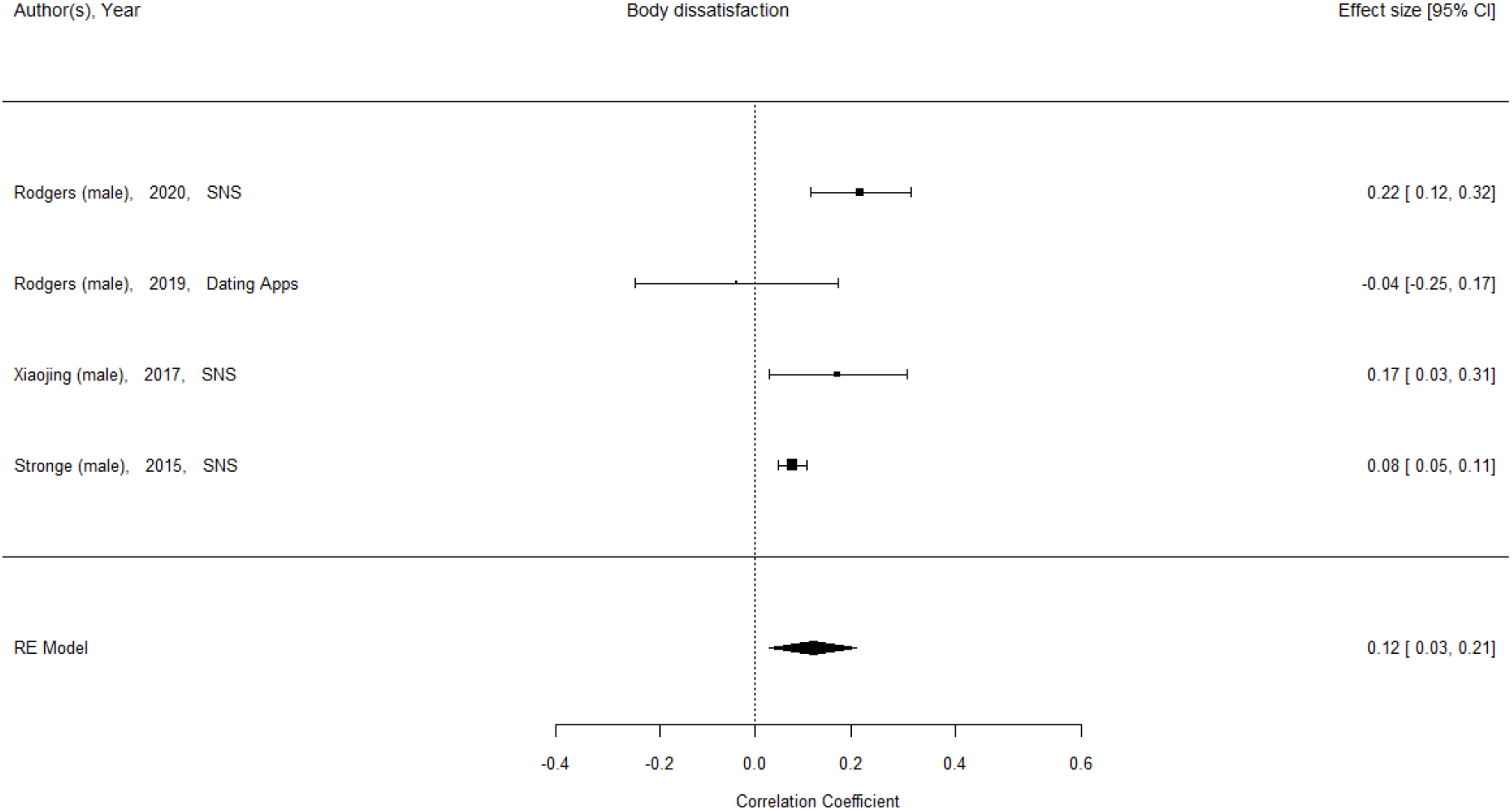
Meta-analysis of Problematic usage of the internet (PUI) by ‘body dissatisfaction’, using Pearson correlation for studies using male samples only. Positive Pearson correlation indicates that a higher score of PUI is associated with higher body dissatisfaction. Correlation values of 0.10, 0.30, and 0.50 represent small, moderate, and large effects, respectively

### 3.4 Drive for thinness

We identified ten studies (n = 3572) reporting data in the drive for thinness domain (Butkowski et al., 2019; Hendrickse et al., 2017; Hummel and Smith, 2015; Kim and Chock, 2015; Kvardova et al., 2020; Levinson et al., 2017; Meier and Gray, 2014; Simpson and Mazzeo, 2017; Tao, 2013; Tiggemann and Miller, 2010). In performing meta-analysis for the drive for thinness domain, we removed one of the studies that was highly influential (reporting an extremely positive Pearson correlation r = 0.51, Cook’s d > 2 S.D. above mean of the cohort of studies, n = 445) and of weak quality (Kvardova et al., 2020). Moreover, this study used ‘web content internalization’ which was a quite dissimilar and appearance oriented PUI metric, which may explain the reported association. By doing so, we report a more conservative pooled estimate. The pooled estimate of the remaining nine studies was small, r = 0.16 (s.e. = 0.037, p< 0.001) (Figure 8).

**Figure 8.**
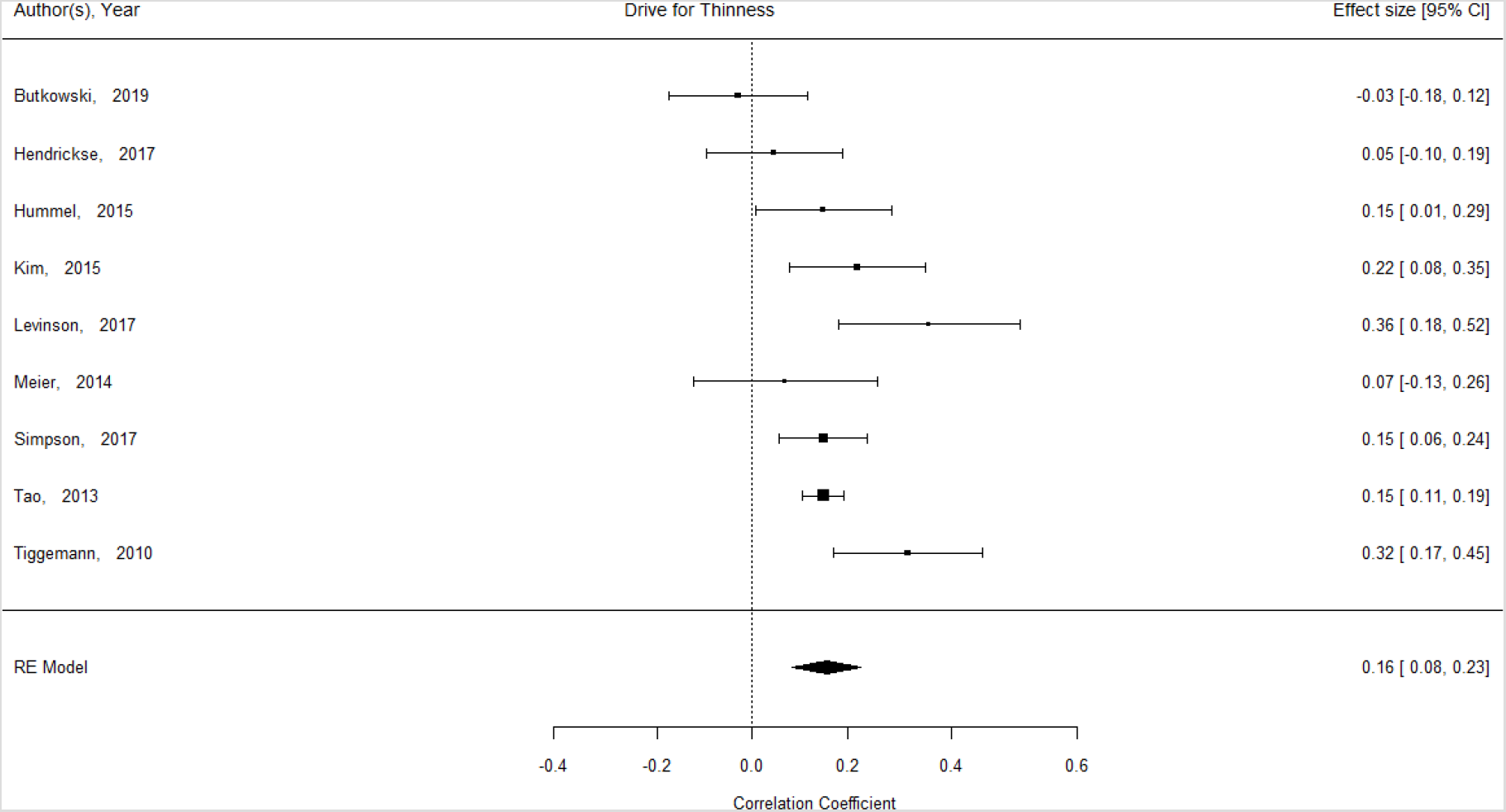
Meta-analysis of Problematic usage of the internet (PUI) by ‘drive for thinness, using Pearson correlation. Positive Pearson correlation indicates that a higher score of PUI is associated with higher drive for thinness. Correlation values of 0.10, 0.30, and 0.50 represent small, moderate, and large effects, respectively

### 3.5 Dietary restraint

Six studies were included in the dietary restraint domain (n = 2397) (Hummel and Smith, 2015; Levinson et al., 2017; McLean et al., 2015; Niu et al., 2020; Rodgers et al., 2020; Simpson and Mazzeo, 2017) providing a small pooled effect estimate, r = 0.18 (s.e. = 0.03, p< 0.001) (Figure 9).

**Figure 9.**
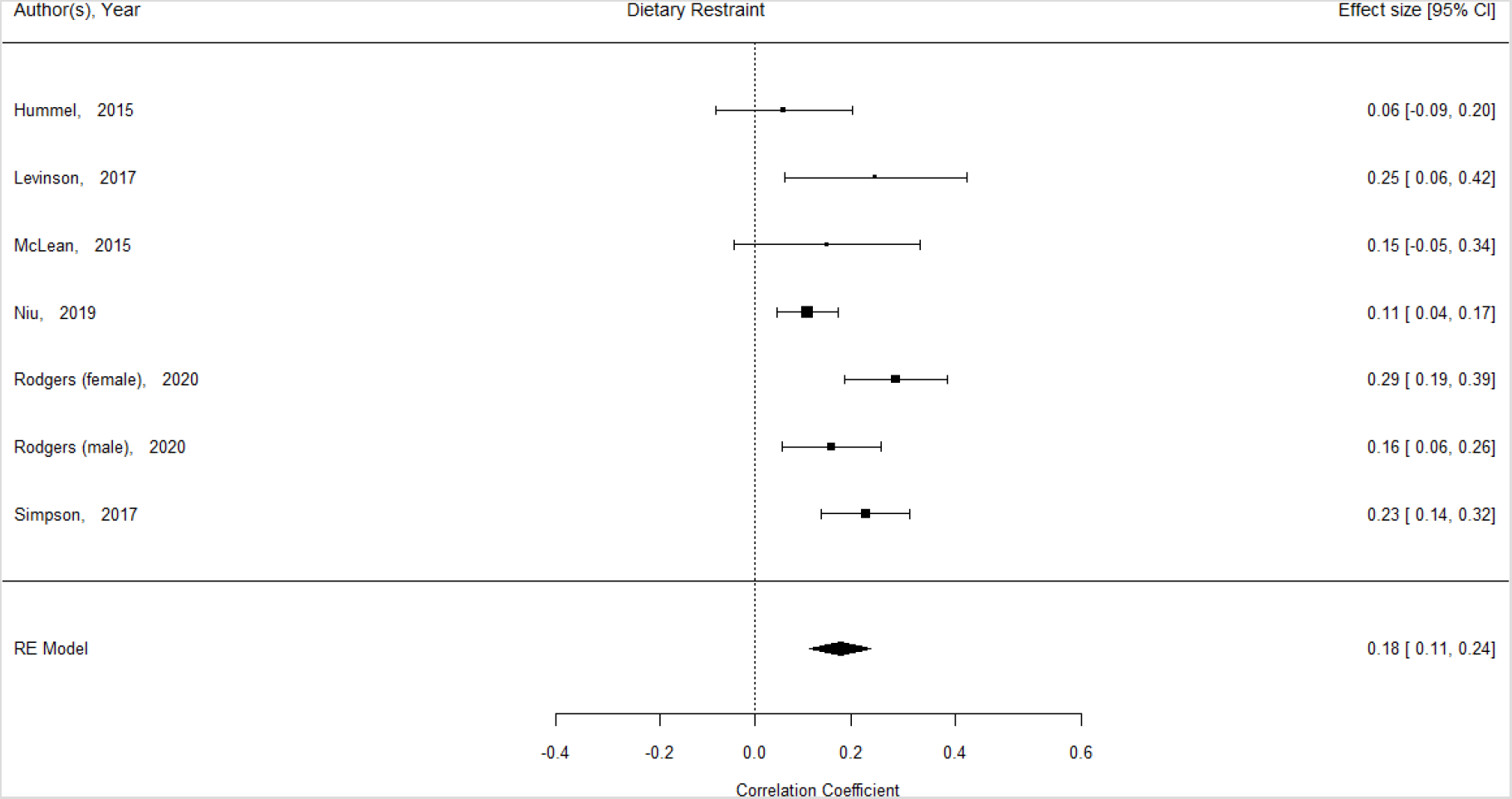
Meta-analysis of Problematic usage of the internet (PUI) by ‘dietary restraint’, using Pearson correlation. Positive Pearson correlation indicates that a higher score of PUI is associated with higher dietary restraint. Correlation values of 0.10, 0.30, and 0.50 represent small, moderate, and large effects, respectively

### 3.6 Moderation analysis

Gender, age group, geographical area of study reporting, PUI facet and quality of study (assessed separately by EPHPP and by our ad-hoc study-specific instrument) were entered in moderation analyses to examine whether they constituted significant moderating factors in any of the effects examined. Results are summarized in Table 2. Gender, geographical area of reporting, PUI facet and study quality (EPHPP or ad-hoc) did not significantly moderate the results in any of the domains examined. Age was found to moderate the effects in the ‘drive for thinness’ domain; (adults > youth, children), moderator all ANOVA comparisons (p< 0.001). Geographical area of reporting was found to moderate the effects in the ‘at-risk eating disorders’ domain; (Africa > Americas, Asia, Oceania and Europe), moderator all ANOVA comparisons p< 0.001).

**Table 2.** Moderation analysis. ***M***eta-analysis was done using random-effects model using REML. ***REML***: Restricted maximum-likelihood estimator; Significances and p-values describe test of moderators in the mixed effects model; ***PUI*** = problematic usage of the internet; ***EPHPP*** = “Quality Assessment Tool for Quantitative Studies” developed by the Effective Public Health Practice Project (EPHPP)

| Domain | Age | Gender | Geographical area of reporting | PUI facet | Quality Ad-hoc | EPHPP |
| --- | --- | --- | --- | --- | --- | --- |
| At-risk eating disorders | n.s. | n.s. | p = 0.019 | n.s. | n.s. | n.s. |
| Body dissatisfaction (female only & mixed) | n.s. | n.s. | n.s. | n.s. | n.s. | n.s. |
| Body dissatisfaction (males only) | n.s. | - | n.s. | n.s. | n.s. | n.s. |
| Drive for thinness | p = 0.042 | n.s. | n.s. | n.s. | n.s. | n.s. |
| Dietary restraint | n.s. | n.s. | n.s. | n.s. | n.s. | n.s. |

### 3.7 Publication bias

We examined funnel plots for all meta-analyses and performed the Regression Test for Funnel Plot Asymmetry in all domains. We did not identify publication bias in any domain (Figure 10).

**Figure 10.**
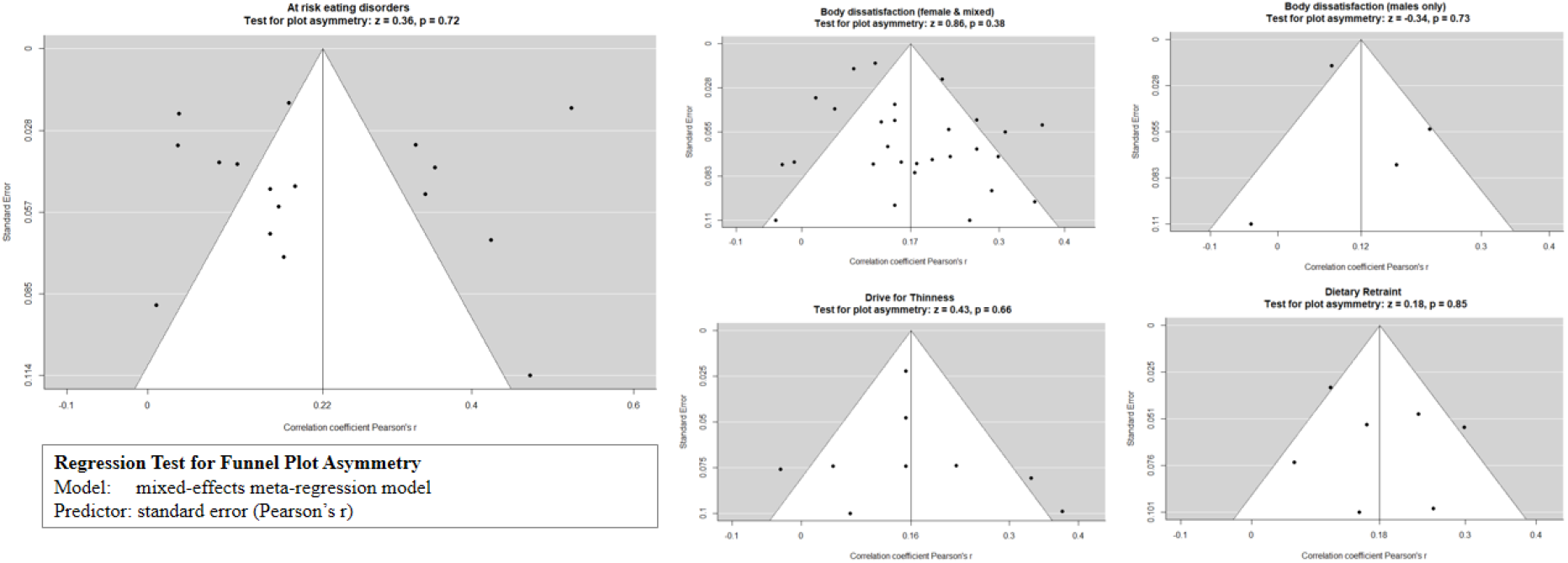
Meta-analysis funnels plots by cognitive domain; ‘z’ and ‘p’ values reported from Regression Test for Funnel Plot Asymmetry (mixed-effects meta-regression model). There was no evidence of publication bias.

## 4 Discussion

This is the first study to amass all quantitative studies under the umbrella of problematic usage of the internet and pool associations on eating disorder and related psychopathology. The meta-analytic results indicate that PUI has a small but significant effect in general ED symptomatology, body dissatisfaction, drive for thinness and dietary restraint. Males experienced higher degrees of body dissatisfaction with higher levels of PUI when examined separately. Moreover, gender did not moderate any of the effect sizes in any domain, neither did quality of study. The systematic review has revealed a prolific research field spanning across the globe and identified unprecedented facets of PUI that are linked to EDs: excessive use of social media, consumption of pro-ED content, overuse of calorie counting and fitness applications, overuse of dating apps and cyberbullying victimization (Figure 11).

**Figure 11.**
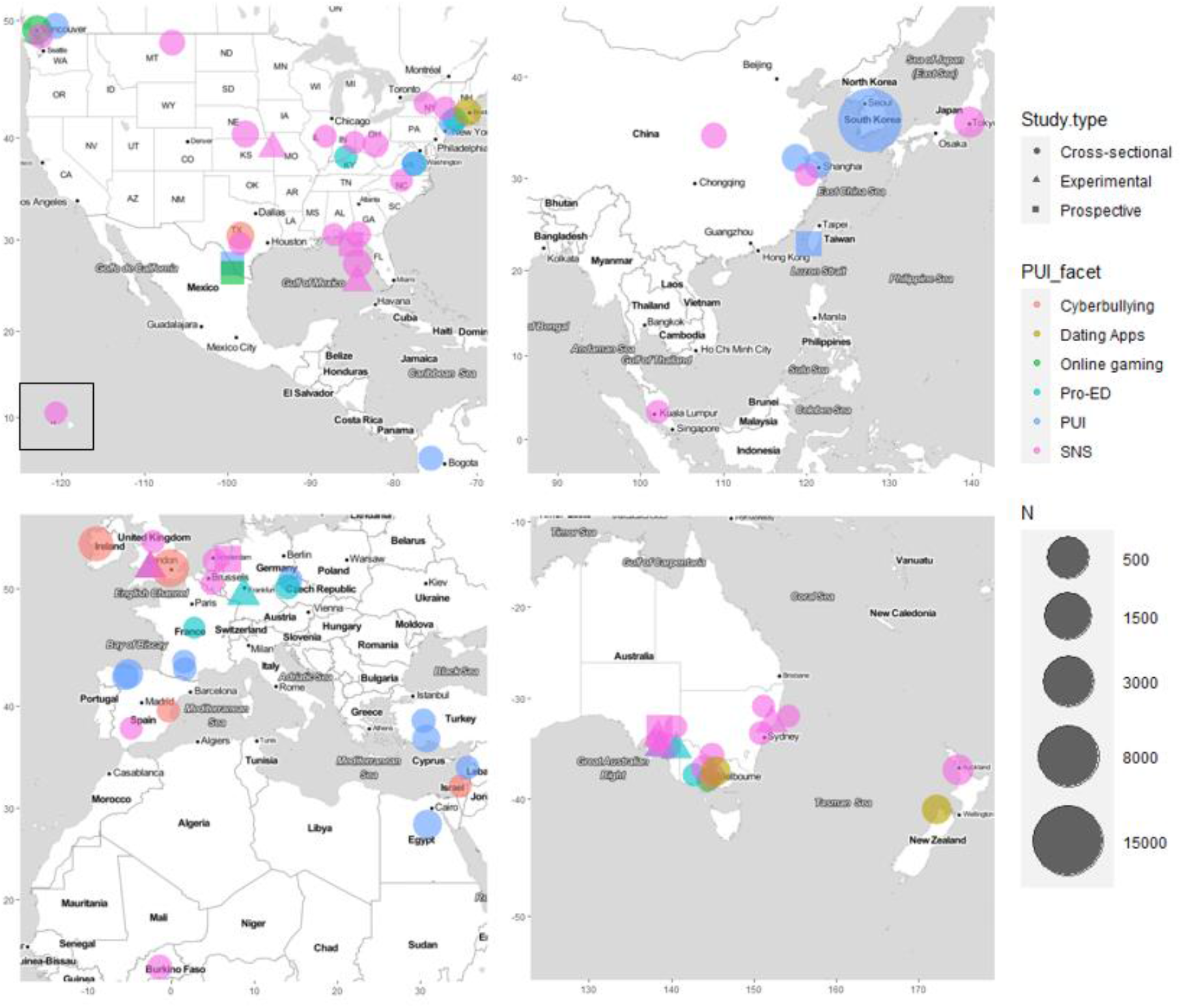
Global map of studies included in the systematic review, indicating the facet of Problematic internet use examined and the size of the study. Geolocation was identified either by the indicated base of where the study took place or alternatively the affiliated address of the first author. Geolocation in areas with high density of studies has been inaccurate on purpose to allow better representation of study density. Studies examining two facets of internet use equally are presented for each facet separately (twice). Hawaii is presented in a separate box together with the Americas.

While our meta-analysis has revealed small but significant associations, it is important to note that those derive from cross-sectional cohort studies that are not designed to explore causality. Even small associations can have significant and clinically meaningful effects if we consider the cumulative exposure or engagement with a particular online activity over a period of time. For example, in one study (Smith et al., 2013), all prospective (T2) associations between PUI and EDI items were stronger as compared to the cross sectional ones (e.g. EDI T1 vs. T2: EDI-Bulimia r = 0.21 vs. 0.26; EDI-BD r = 0.20 vs. 0.26; EDI shape concern r = 0.26 vs. 0.30). Similar findings were described in another study (Hummel and Smith, 2015), but not consistently across all EDI subscales and PUI metrics. In our systematic review, we identified a number of prospective and experimental studies that aimed to disentangle those associations from a causational point of view. While those studies did not qualify for inclusion in the meta-analysis, they are a very important complement in our understanding of the main results and are discussed below. We also discuss in more detail specific facets of ED psychopathology (e.g. bulimia symptoms) and PUI (internet gaming, SNS usage, dating Apps, cyberbullying victimization) for which substantial literature exists as an important complement to the meta-analysis results.

### 4.1 Prospective studies

There are a number of prospective observational studies in the field: Smith et al. followed up a cohort for four weeks (n = 219, USA) and found that SNS usage increased bulimic symptoms and episodes of over-eating (EDI-BN) in a non-clinical sample of female college students (Smith et al., 2013). In a similar study, Hummel et al. (Hummel and Smith, 2015) followed up a similar student cohort for four weeks (n = 177, USA) and showed that receiving negative feedback through Facebook interactions can negatively impact on eating attitudes. Heavier Facebook usage led to stronger impact of association. De Vries et al., followed up a cohort of adolescents (n = 604, native Dutch) and showed that SNS use predicted increased body dissatisfaction; those results were not moderated by gender (de Vries et al., 2016), while Ferguson et al. followed up a cohort of adolescent females (n = 101, USA) for six months and found that mood symptoms may be mediating the effects of SNS on ED symptoms concurrently and prospectively (Ferguson et al., 2014). Tiggeman et al. followed up a cohort of high school children for two years (n = 438, South Australia); the number of Facebook friends predicted DFT two years later, while internalization and body surveillance predicted an increase in the number of friends (Tiggemann and Slater, 2017). Those results are in support of the notion that cumulative exposure to PUI over time has a role in the fuelling eating disorder pre-disposing factors and psychopathology. On the other hand, Hsieh et al. followed up a student cohort (n = 500, Taiwan) for a year (Hsieh et al., 2018) to identify predictors of development of and remission from Internet Addiction (IA). They examined bingeing and purging behaviors (by single-item question) as potential parameters; neither was associated with the development of or remission from IA, therefore, although the ascertainment of ED pathology was limited in this study, a ‘reverse’ association was not observed.

### 4.2 Experimental studies

Experimental studies have explored the relationship between internet usage and eating disorder symptomatology using various manipulation paradigms: in children, appearance focused internet gaming vs. control (ED neutral) gaming was associated with higher body dissatisfaction (Experimental condition of PUI ∼ BD, d =.46) (Slater et al., 2017). In adult participants, those with elevated risk for developing an ED had a lower appearance self-esteem after exposure to the pro-ED website, compared to neutral condition (Theis et al., 2012); similarly, low mood after SNS exposure was more prominent to those having appearance comparison traits (Fardouly et al., 2015). Furthermore, being allocated to use Facebook 20 min/day was associated with the maintenance of weight/shape concerns as compared to an alternate (ED neutral) internet activity (Mabe et al., 2014). Also, exposure to fitspiration images (Prichard et al., 2020; Tiggemann and Zaccardo, 2015), or celebrity or peer Instagram sourced images (Brown and Tiggemann, 2016) was significantly associated with higher BD relative to exposure to travel (ED neutral) images. On a separate vein, exposure to humorous, parody images of thin-ideal celebrities was associated with higher body satisfaction, versus the actual thin-ideal images alone (Slater et al., 2019); and not moderated by trait levels of ITI, while self-disclaimer captions on images did not alter body dissatisfaction influences (Livingston et al., 2020). Finally, with a different type of experimental study protocol (using ecological momentary assessment), SNS visits were predictive of body dissatisfaction in a group of college students (Bennett et al., 2019). In sum, appearance-focused online gaming and ‘general’ SNS usage (e.g. Facebook exposure) or ‘ED-content’ specific (e.g. consumption of ‘fitspiration’) may have direct effects on body dissatisfaction. Future research may benefit from examining less examined PUI facets or how those effects change in respect to accumulative exposure over time and neurodevelopmental window of exposure (e.g. from childhood to adolescence or early adulthood), as well as consider putative online-based resilience focused interventions to promote body satisfaction via SNS.

### 4.3 Bulimia nervosa

There were too few studies to quantify a pooled estimate for bulimia nervosa symptoms, however, three studies reported Pearson correlations between PUI and bulimia nervosa symptoms, with small to medium correlations (r range [0.11–0.36] and Spearman’s rho = 0.13 (Melioli et al., 2015)) (Butkowski et al., 2019; Smith et al., 2013; Tao, 2013) and statistically significant group differences (Holland and Tiggemann, 2017; Tao and Liu, 2009; Wilksch et al., 2020). Those results appear in line with the general eating disorder psychopathology effects of PUI which are statistically significant and of small to medium magnitude.

### 4.4 Internet gaming

Internet gaming disorder is now a recognized disorder in both DSM-5 and ICD-11 (Jo et al., 2019). Appearance focused gaming may influence body satisfaction in children (Slater et al., 2017); similarly, excess use of internet including gaming was associated with BD in adults (Carter et al., 2017). However, in another study of pre-adolescent girls, SNS use including use of social gaming sites was not related to BD or EAT-26 scores (Ferguson et al., 2014). Therefore, it is still unclear how and what aspect of internet gaming relates to eating disorders.

### 4.5 Social networking sites use

The effect of SNS use on ED and related psychopathology e.g. body dissatisfaction, has been examined in multiple studies that were included in the quantitative synthesis of this paper and beyond those (Fardouly et al., 2015; Fardouly and Vartanian, 2015; Hendrickse et al., 2017; Holland and Tiggemann, 2017; Kim and Chock, 2015; Lee et al., 2014; Sidani et al., 2016; Wagner et al., 2016). Socio-cultural attitudes towards own appearance, comparisons between users and the ITI (Cohen and Blaszczynski, 2015; McLean et al., 2015; Meier and Gray, 2014; Melioli et al., 2015; Tiggemann and Miller, 2010; Tiggemann and Slater, 2017; Xiaojing, 2017), objectified body consciousness (Manago et al., 2015; Tiggemann and Miller, 2010; Tiggemann and Slater, 2013), sexualization and self-objectification (Karsay et al., 2018) as well as self-esteem (Ahadzadeh et al., 2017) appear to be important influencing factors interacting with body image and ED symptoms. ‘SNS addiction’, as a separate entity, has been associated with higher levels of ED psychopathology (Aparicio-Martinez et al., 2019). SNS use has been the most examined facet of PUI in relation to ED problems. Arguably it is confounded with the consumption of pro-ED content, appearance related gaming, dating and cyberbullying victimization which to a degree may be contributing to the observed effects; future research could potentially examined those parameters separately in order to disentangle those associations. This might help shaping the SNSs of the future, by detracting from a potentially blanket demonization of SNS use, and rather regulating or modifying potentially hazardous content and promoting the development of, and engagement in resilience building SNS content.

### 4.6 Dating Apps

A few research studies have examined the association between dating apps and eating disorders (Griffiths et al., 2018b; Rodgers et al., 2019; Tran et al., 2019). Use of dating Apps has been linked with unhealthy weight control behaviors in adults (OR = 2.7—16.2), as well as multiple unhealthy weight control behaviors (laxative misuse, muscle building supplements, self-induced vomiting, diet pill use) (Tran et al., 2019), body shame (Rodgers et al., 2019) and muscularity dissatisfaction and use of anabolic steroids (r = 0.10 and r = 0.09 respectively) (Griffiths et al., 2018b).

### 4.7 Calorie tracking Apps, fitness Apps and consumption of nutrition, weight loss and fitness websites

A number of observational studies have examined weight loss and fitness Apps in conjuncture with body image and eating disorder difficulties (Almenara et al., 2019; Embacher Martin et al., 2018; Simpson and Mazzeo, 2017). Obsessional thinking around weight and activity goals may become attractors of unhealthy behaviors. The use of calorie tracking Apps was associated with body dissatisfaction among college students (Embacher Martin et al., 2018), whereas use of calorie or fitness Apps accounted for more than half of the variance in the degree of ED psychopathology (Simpson and Mazzeo, 2017). In another study, eating disorder psychopathology was associated with visits to weight loss websites (Almenara et al., 2019), whereas following Instagram ‘Health and Fitness’ accounts was moderately correlated with drive for thinness (Cohen et al., 2017). In a male only cohort, EDE-Q scores were much higher among fitness App users (n = 122, Australia) (Linardon and Messer, 2019), whereas in females the use of My Fitness Pal was correlated significantly with degree of ED psychopathology (Levinson et al., 2017). Finally, self-reported eating disorder was associated with consumption of fitness, weight loss and fitspiration content (Carrotte et al., 2015). It is important to note that all of those studies were of cross-sectional online survey design, with various methodological limitations. Overall, the field could benefit from further investigations on calorie tracking and fitness Apps or other online media that specifically support weight loss and their relationship with eating disorders.

### 4.8 Cyberbullying victimization and eating disorders

There were not enough studies to allow separate meta-analysis on this PUI facet using quantitative data (Kelly et al., 2018; Kenny et al., 2018; Marco et al., 2018; Olenik-Shemesh and Heiman, 2017; Pistella et al., 2019). However, our narrative synthesis identified profound effect between the prevalence of cyberbullying and how one perceives their body, leading to disordered eating patterns and negative body appraisal. The majority of research suggests that women are much more likely to become a victim. It has been found that girls and women could be as much as three times as likely to recall their body as “too fat”, if they experienced cyberbullying, as opposed to males (Kenny et al., 2018; Marco et al., 2018; Olenik-Shemesh and Heiman, 2017). Cyberbullying victimization has been reported to have debilitating mental health side effects (Kelly et al., 2018), such as poor sleep, low self-esteem and body dissatisfaction, all of which has also been proven to lead to negative body perception (Griffiths et al., 2018a). These factors are mediated, however, by the length of time that is spent on social media.

### 4.9 Limitations

In respect to our quantitative analysis, there are limitations to consider deriving from the heterogeneity of PUI measurements, as well as from the heterogeneity in the eating disorder domains of ascertainment.

In our selected studies, we identified a paucity of exploration of confounding comorbidities; less than 10% of the studies considered mood and anxiety disorders and less than 5% considered attention deficit and hyperactivity problems, impulse control difficulties, substance abuse or other psychopathology. Those are well documented comorbidities of PUI (Chamberlain et al., 2018; Fineberg et al., 2018; Ho et al., 2014; Ioannidis et al., 2016) and should be taken into consideration when examining causal effects. For example, at risk eating disorders and PUI subgroups we associated with higher degrees of depression and impulsivity, whereas the co-occurrence of PUI and ED was associated with greater depression when PUI or ED was considered separately (Ivezaj et al., 2017). However, in the largest study of this kind (n = 70,696, Korea) (Park and Lee, 2017), internet addiction was associated with problematic weight control behaviors, before and after adjusting for confounders (e.g. sleep satisfaction stress, depressive mood). Overall, the lack of detailed putatively confounding comorbidities in those studies requires that any causal link should be drawn cautiously.

We included body-dysmorphic disorder (BDD) in our inclusion strategy, as individuals with BDD also present with body dissatisfaction similar to those with eating disorders (Hrabosky et al., 2009). BDD may be confounding some of the observed relationships and careful examination of this diagnostic entity is warranted. However, our search identified very few papers that have examined BDD and internet usage and none considered this as a confounder of eating disorder psychopathology. More research is required to understand the relationship between PUI and BDD, exercise addiction (Corazza et al., 2019) and the internalization of muscular ideals (Rodgers et al., 2020). Those at risk of BDD and exercise addiction are also more inclined to use Image and Performance Enhancing Drugs (IPEDs), including a wide range of untested fitness/weight loss supplements often used without clinical supervision (Corazza et al., 2019; Corazza and Roman-Urrestarazu, 2019; Mooney et al., 2017).

Similarly, we did not identify enough studies to comprise a meaningful quantitative comparison or narrative synthesis for exercise addiction, “mukbang” (an online audiovisual broadcast in which a host consumes large quantities of food while interacting with the audience) in relation to binge eating, cyberchondria (or digital health anxiety) in relation to healthy eating e.g. “orthorexia”. Future studies may be able to describe and ascertain those relationships in greater level of detail. Another related concept, ‘desire for slimness’ has been explored in a large cohort of children (n = 4330, Japan) (Sugimoto et al., 2020); SNS was associated with desire for slimness in girls but not in boys. Further research is required to ascertain if there is any causal link with the development of eating disorder psychopathology later in life.

While we did not find any notable moderating effects from geographical area of reporting, we did not find adequate reporting of race or ethnicity and race/ethnicity stratified metrics to ascertain meta-analytically whether there exist any moderating effects. Ethnic background, including cultural ideals and biological race might make some individuals less susceptible to the effects of PUI on eating disorders psychopathology or body dissatisfaction (Awad et al., 2015; Molloy and Herzberger, 1998). Unfortunately, just more than half of included studies reported detailed ethnicity data and the vast majority did not utilize those in meaningful stratified analysis.

## 5 Conclusion

We have shown that PUI is associated with statistically significant associations with eating disorder psychopathology, body dissatisfaction, drive for thinness and dietary restraint, with results putatively expanding to bulimia nervosa symptoms. Males experience similar degree of body dissatisfaction. Multiple facets of PUI seem important to understand the effects of PUI on the development and maintenance of eating disorders, including excessive consumption of SNS and pro-ED content, overuse of calorie tracking and fitness Apps, overuse of dating apps and experiencing cyberbullying victimization. This provides crucial insight into the importance of online behaviors towards the development and maintenance of eating disorders, as well as our understanding of PUI as a multifaceted concept.

## Data Availability

Data for this manuscript can be provided by the corresponding author upon reasonable request

## 7 Author contributions

KI designed the idea for the manuscript, analyzed the data, wrote the majority of the manuscript and coordinated the co-authors’ contributions. Investigators KI, CT and KB performed inclusion/exclusion processing; KI, ARU, CT and LH independently extracted data from full texts and performed qualitative analysis. All authors read and approved the final manuscript and contributed to the drafting and revising of the paper as well as to interpreting the results.

## Acknowledgement

We are indebted to the authors who kindly provided data for meta-analysis upon request. The authors would like to acknowledge the European Cooperation in Science and Technology COST Action 16207: European Network for Problematic Usage of the internet www.cost.eu. The views presented in this manuscript are not necessarily those of COST.

---

**TABLE S1.**
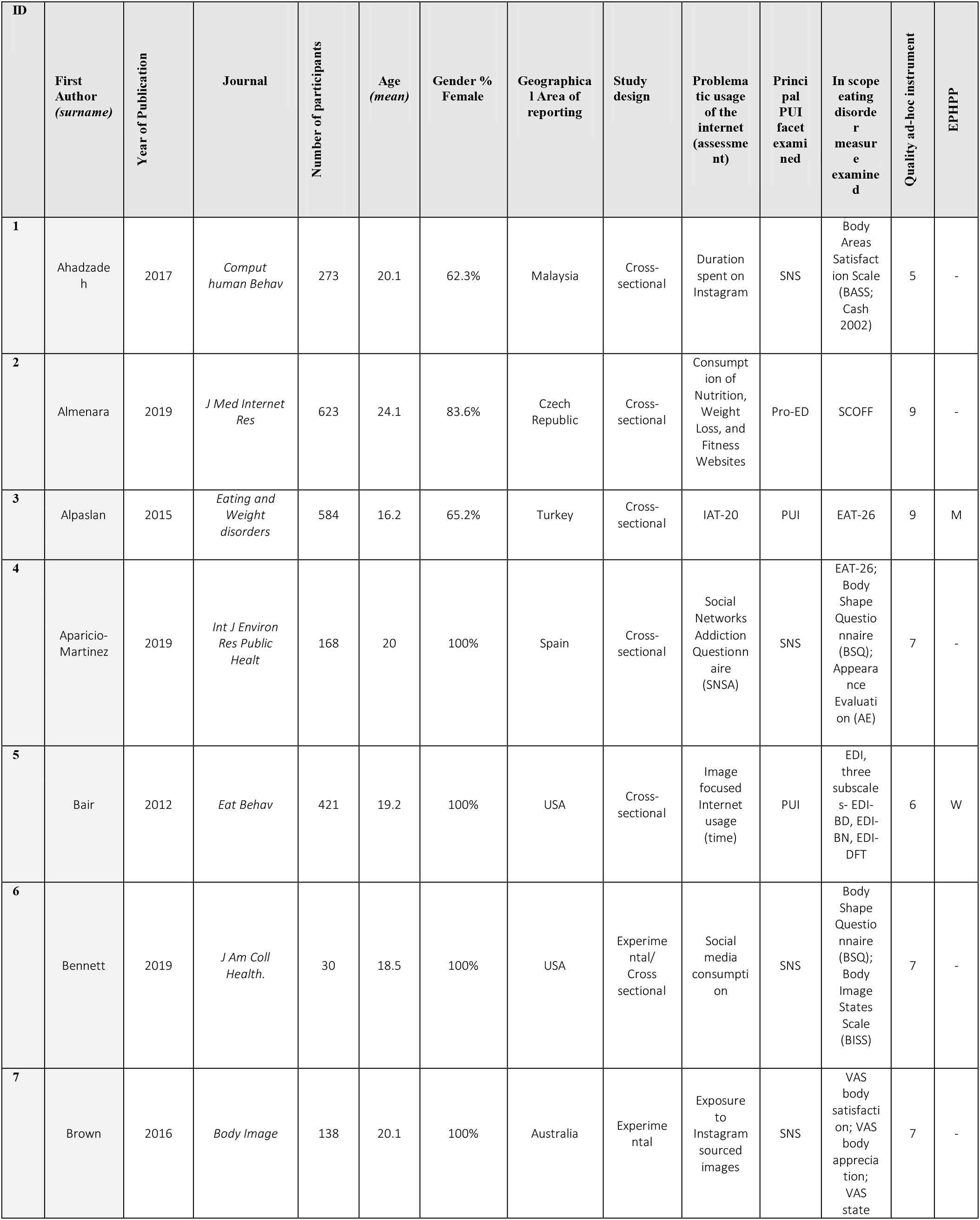
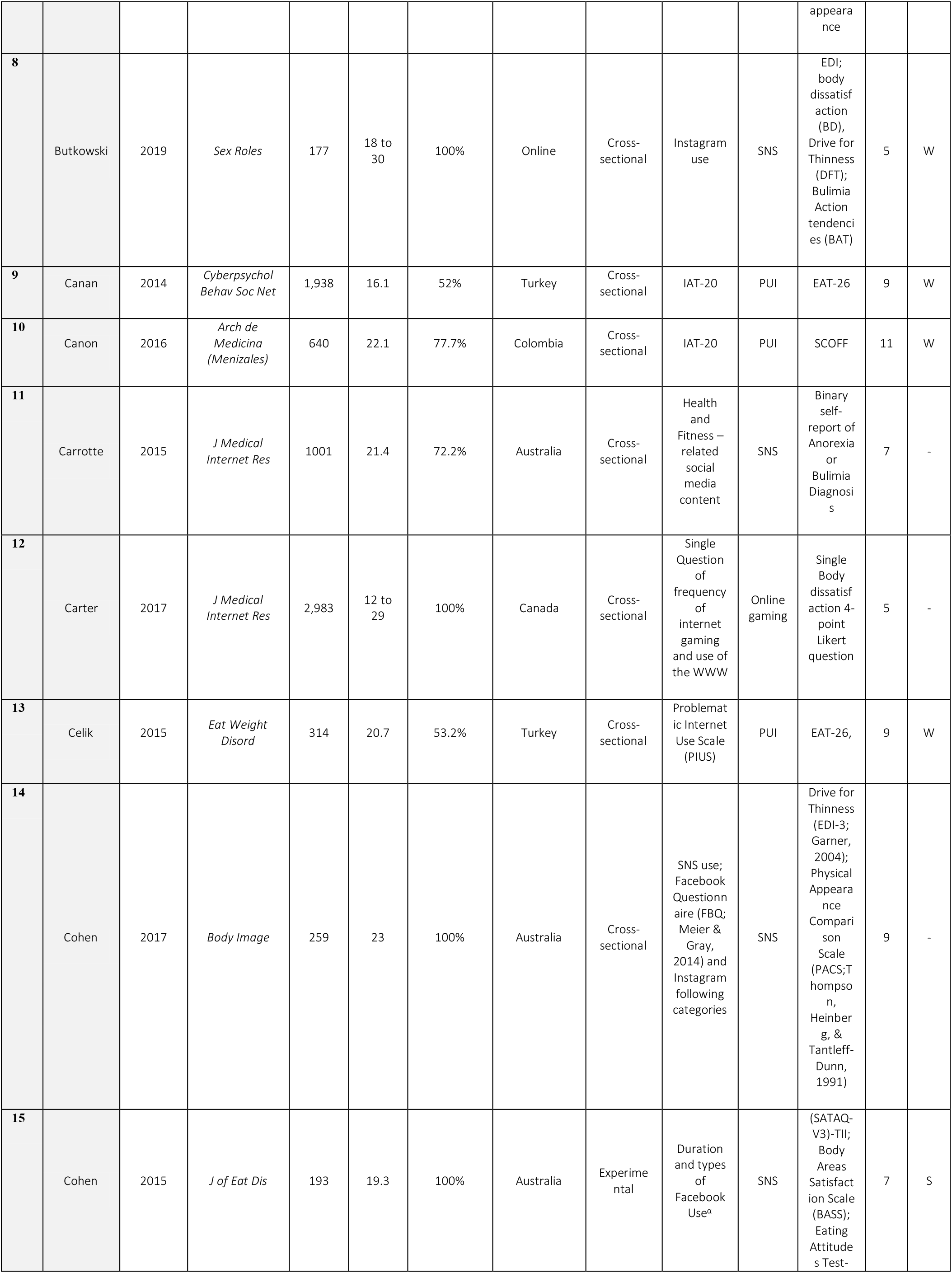
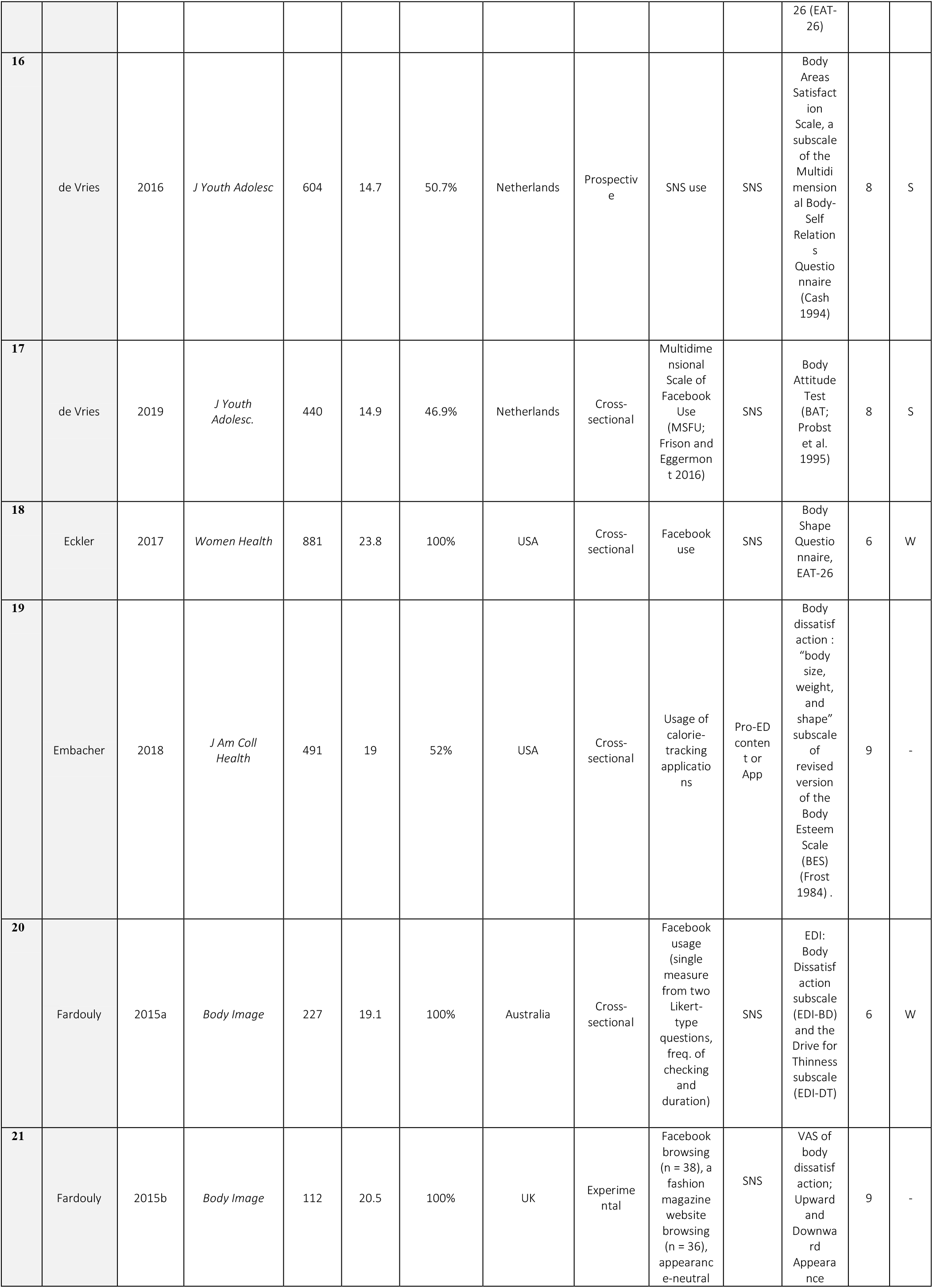
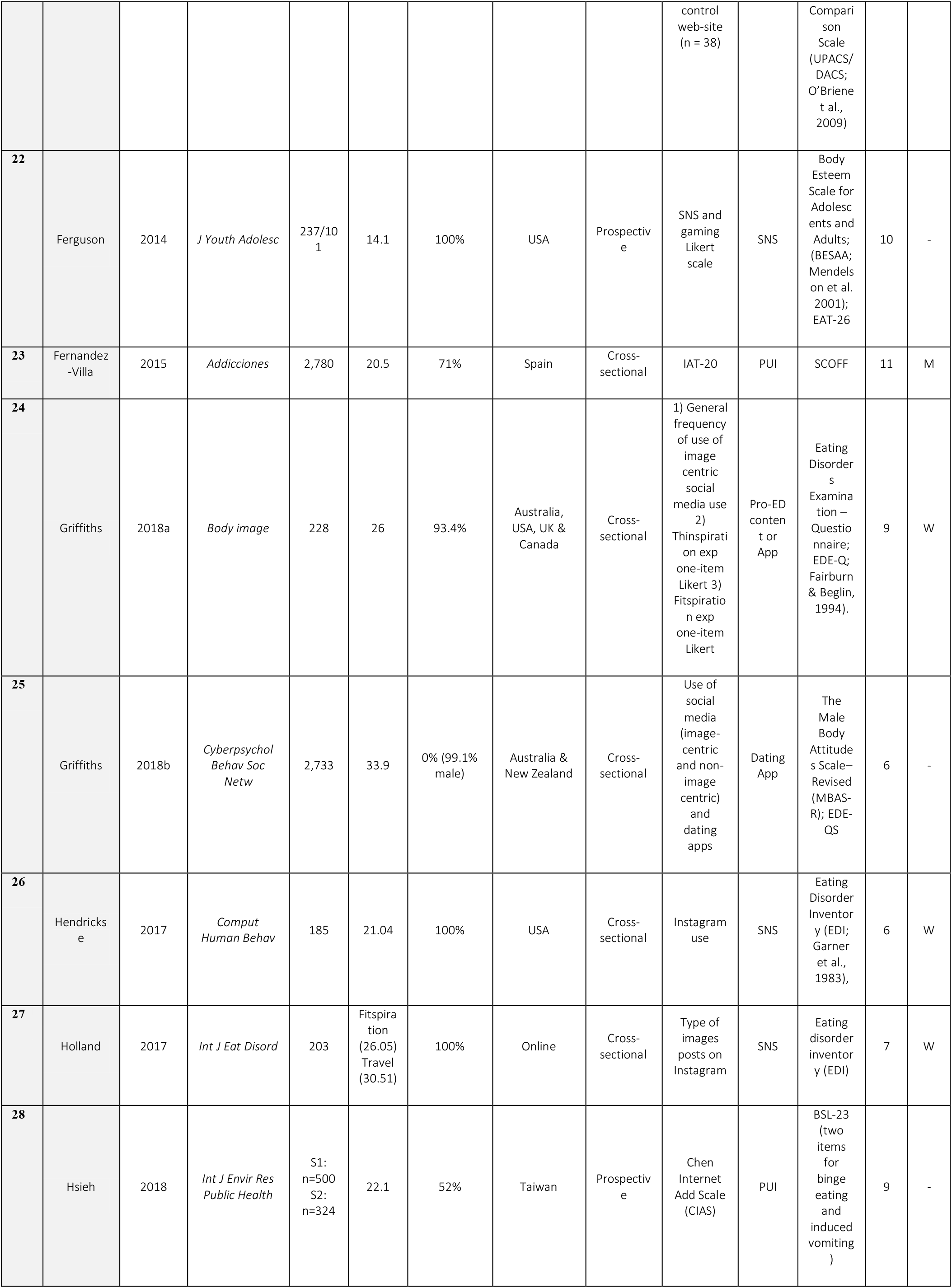
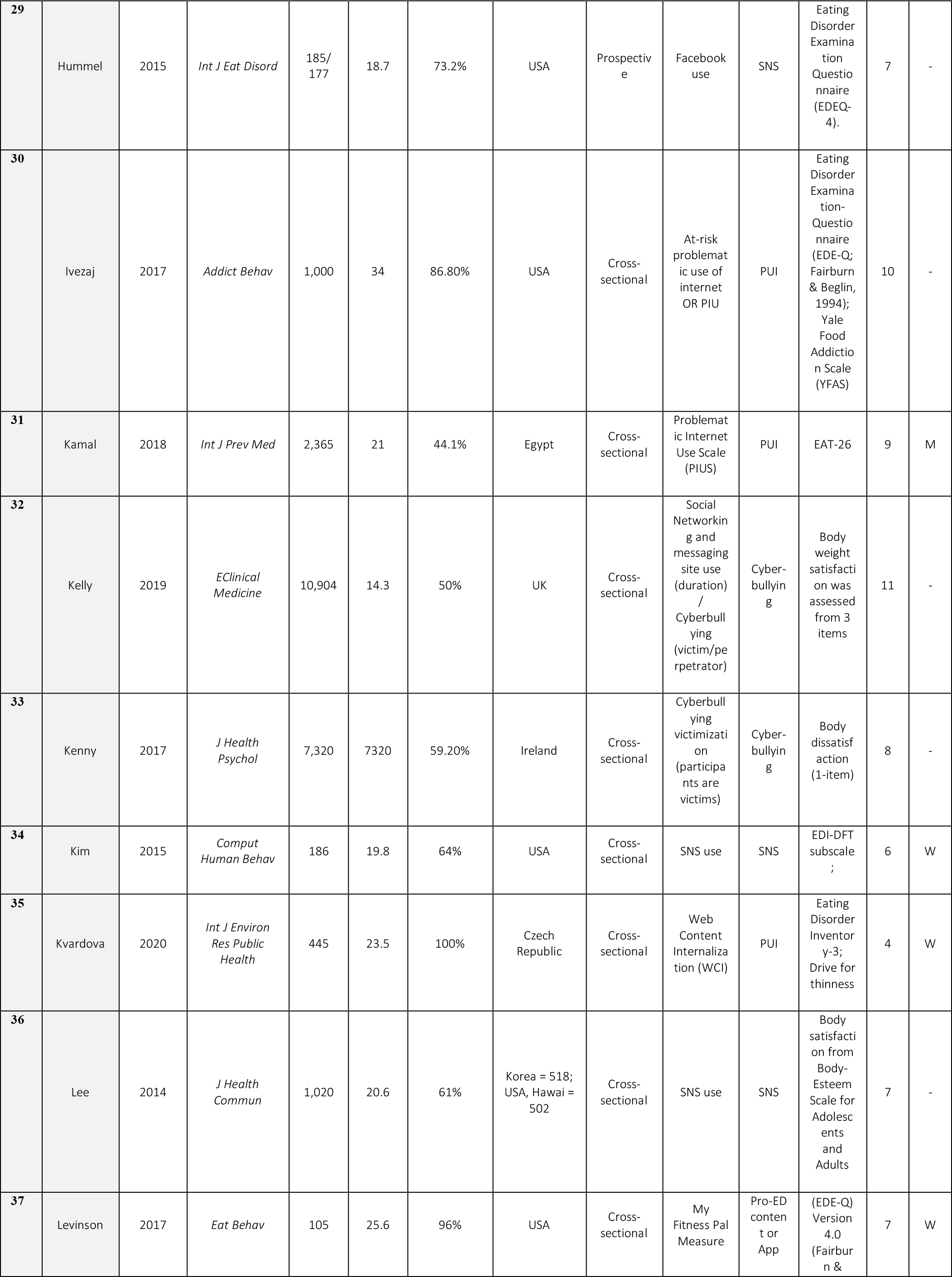
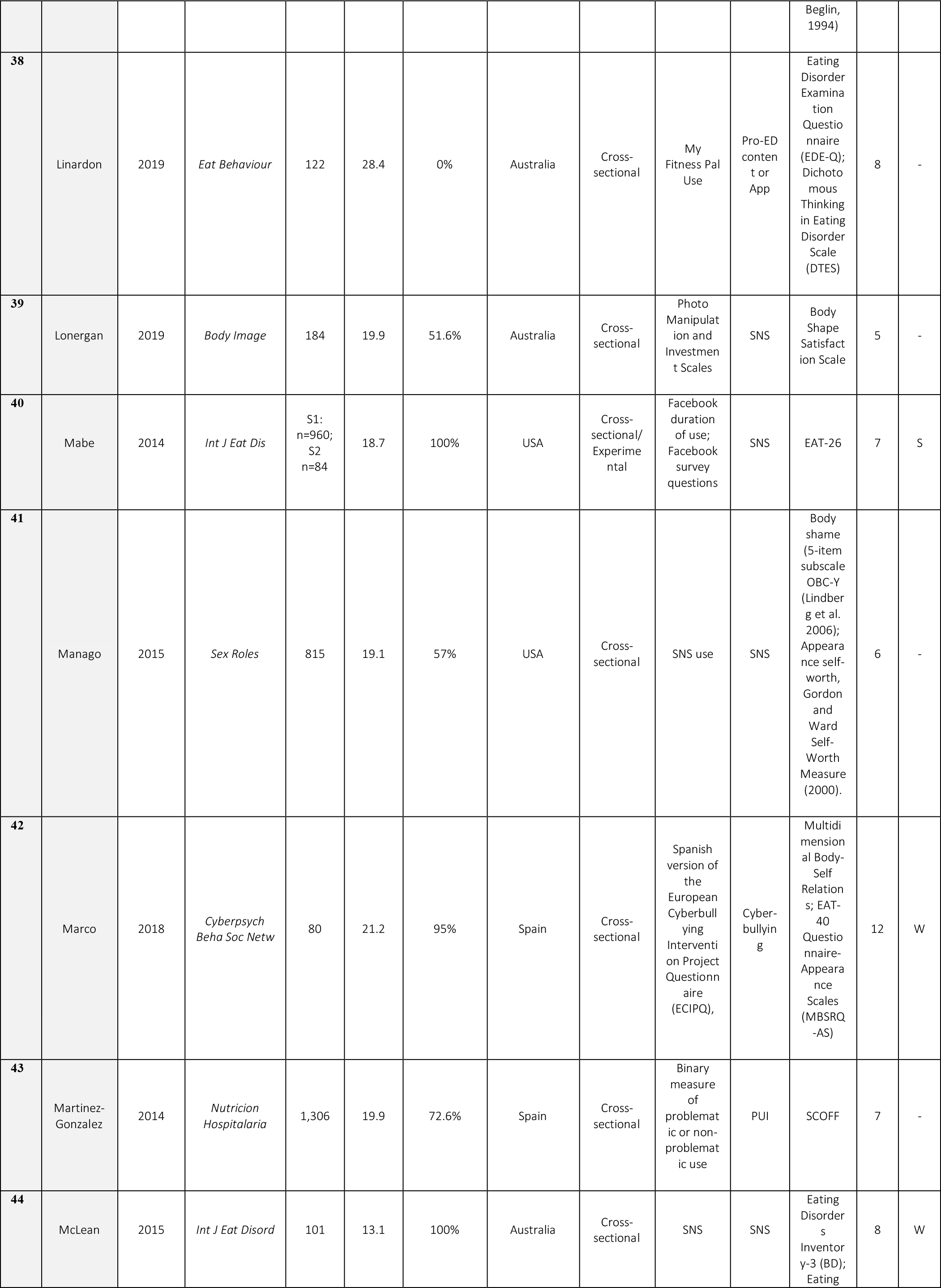
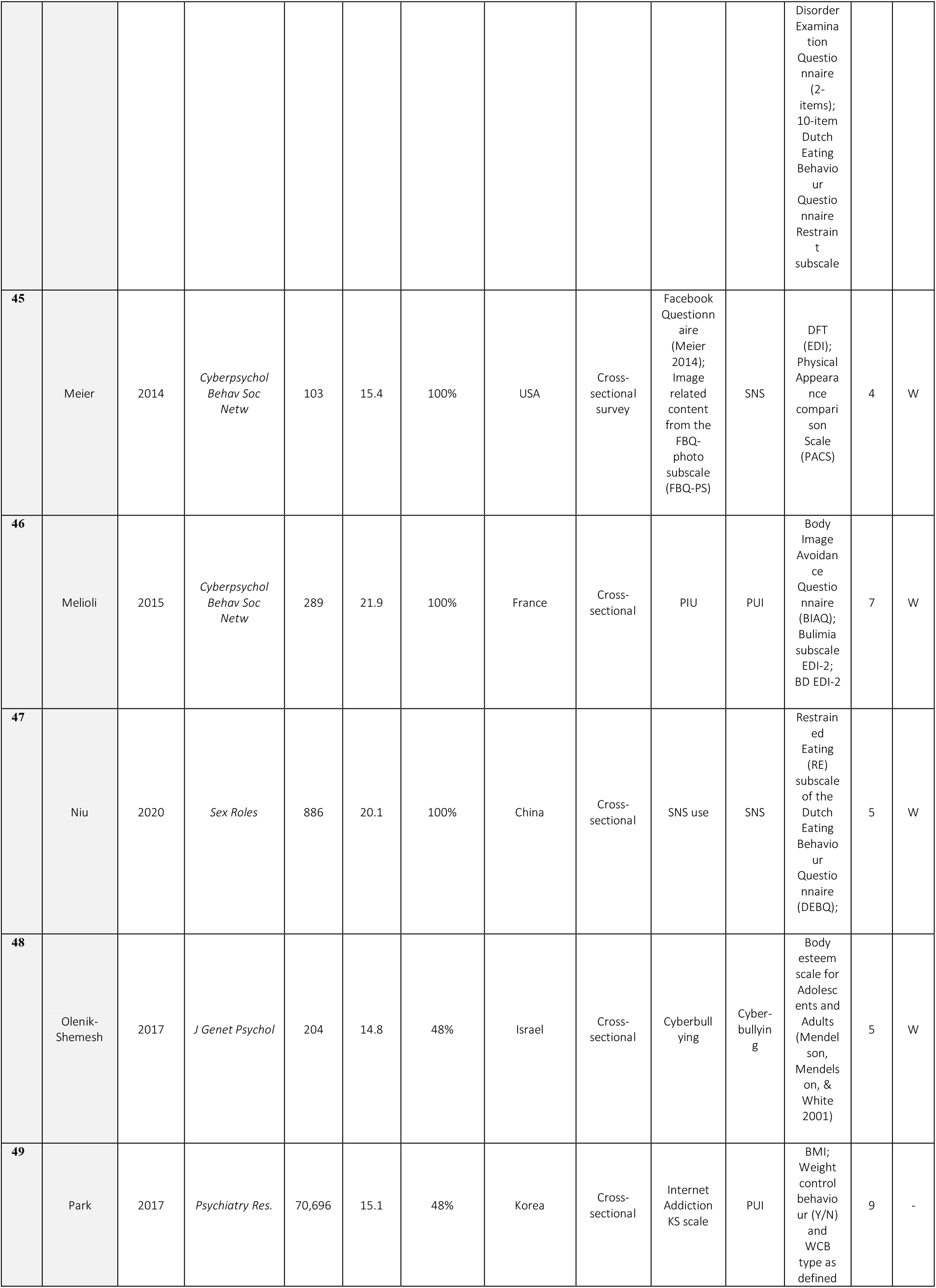
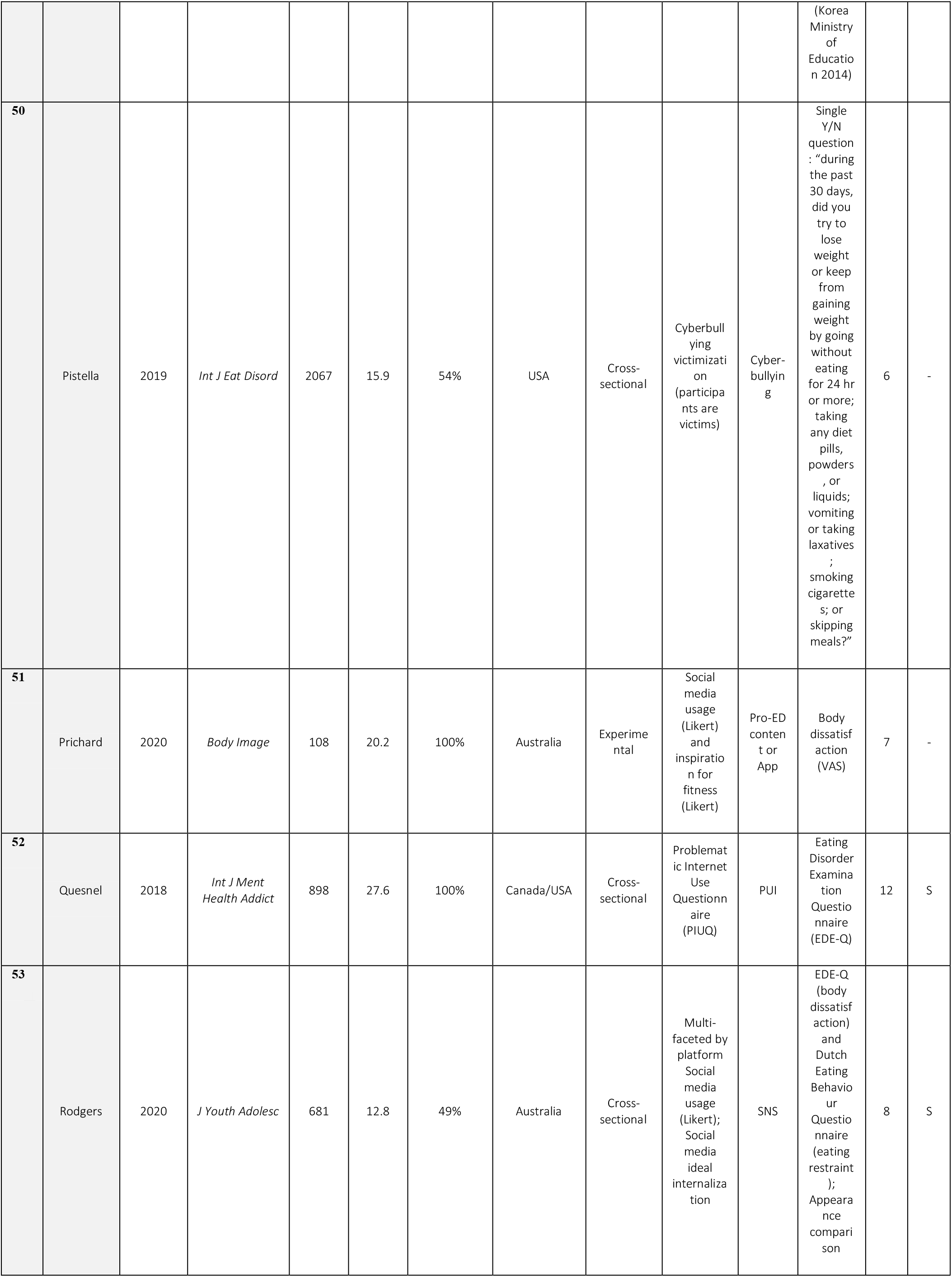
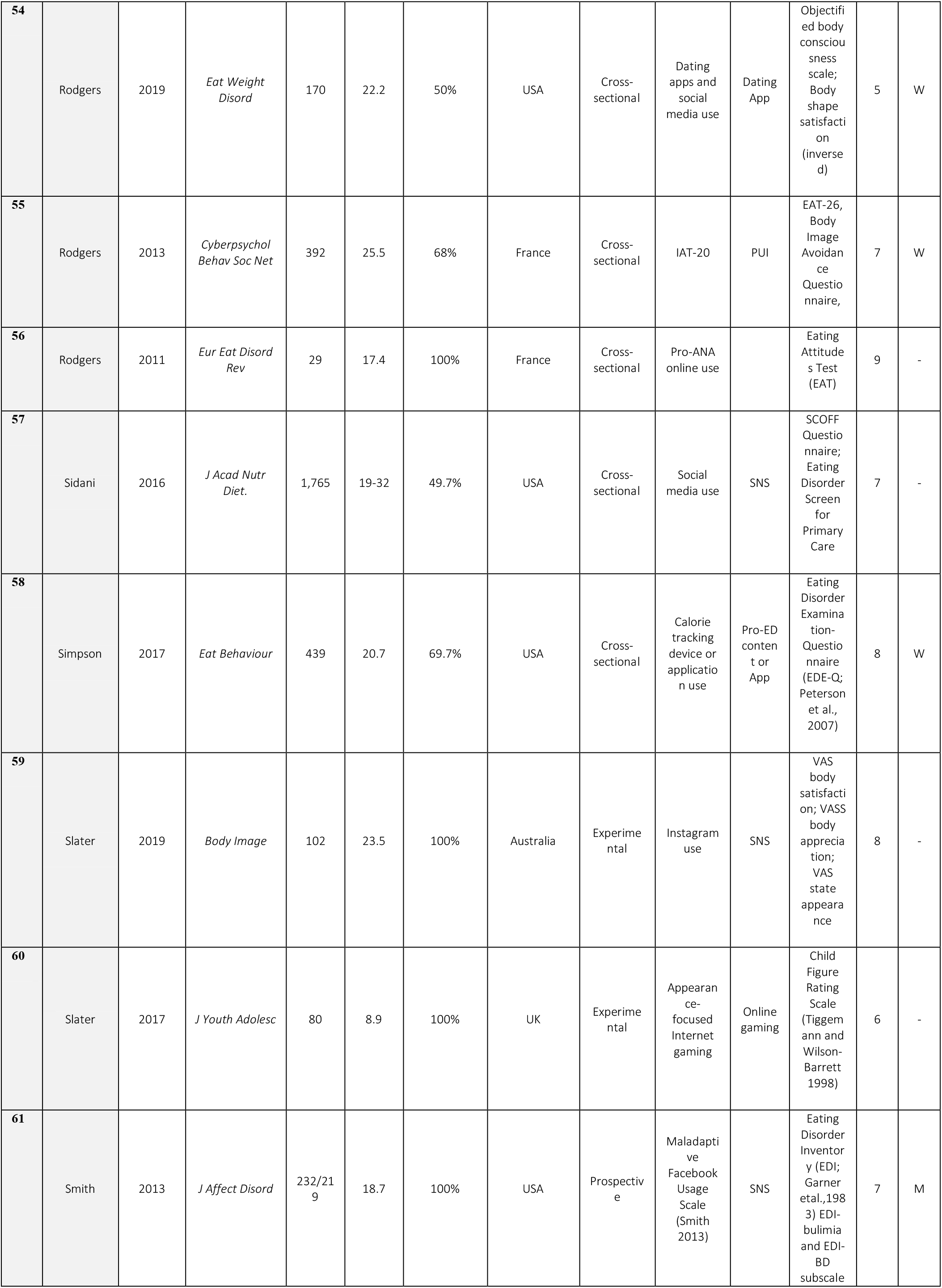
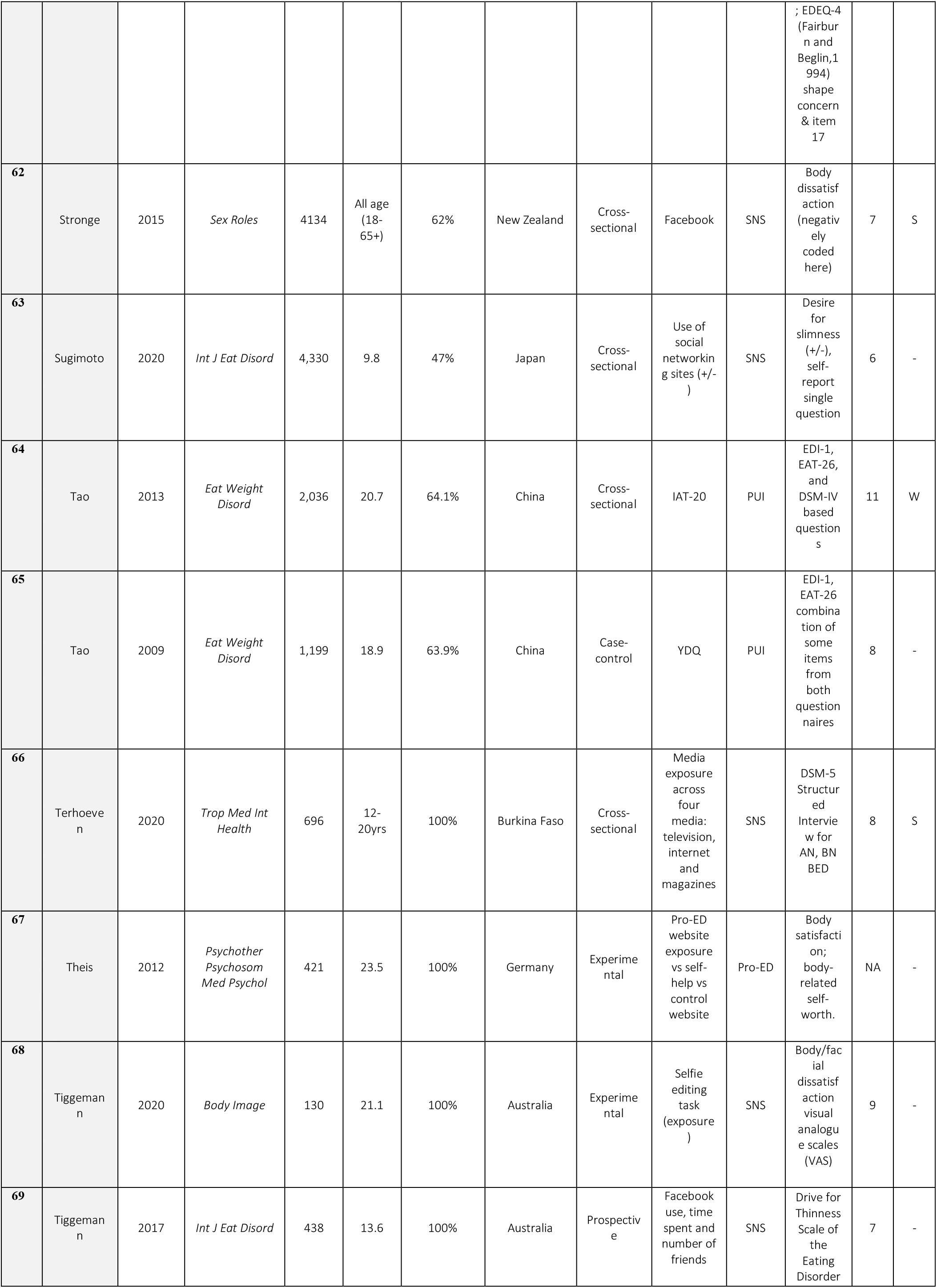
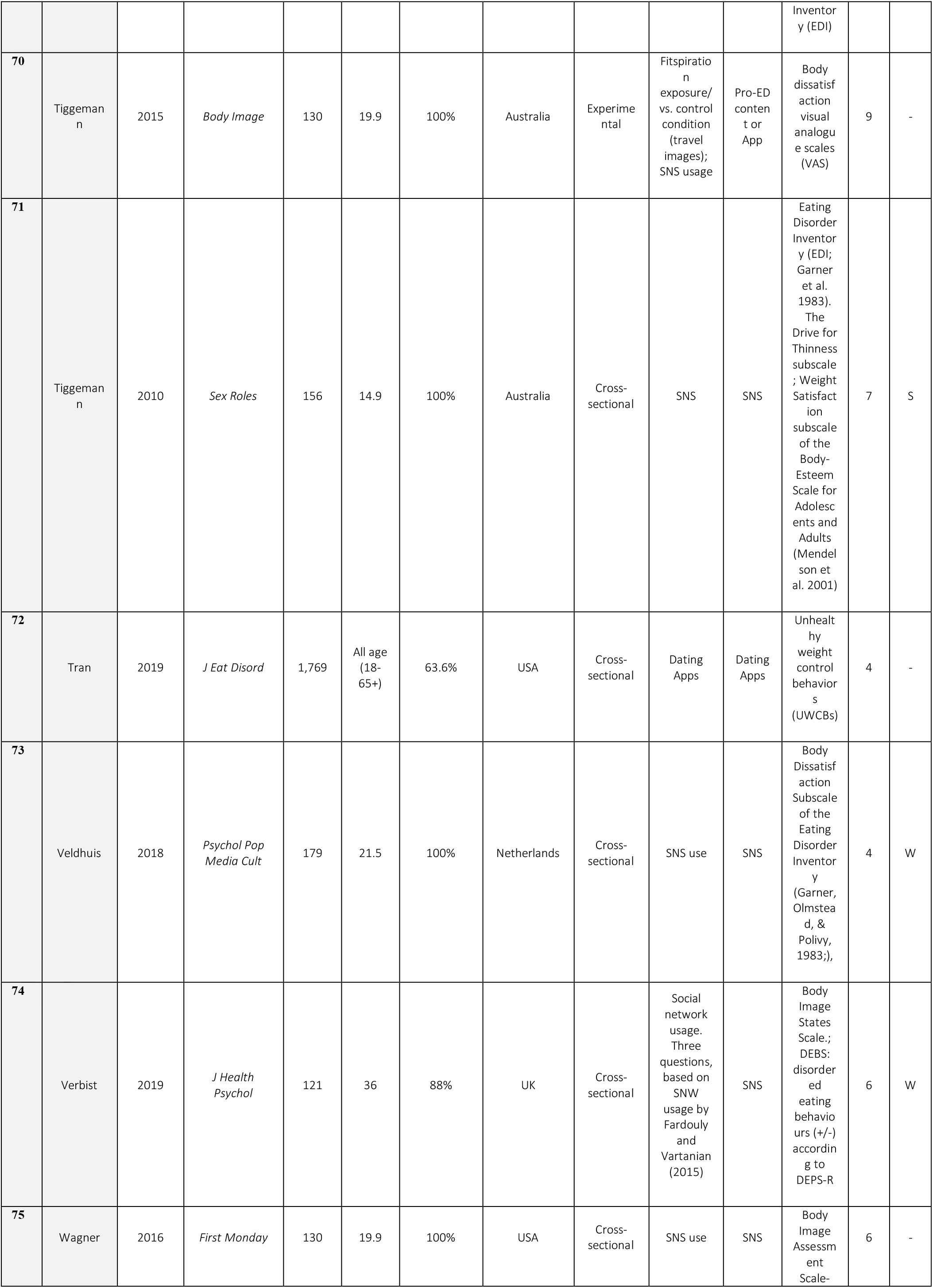
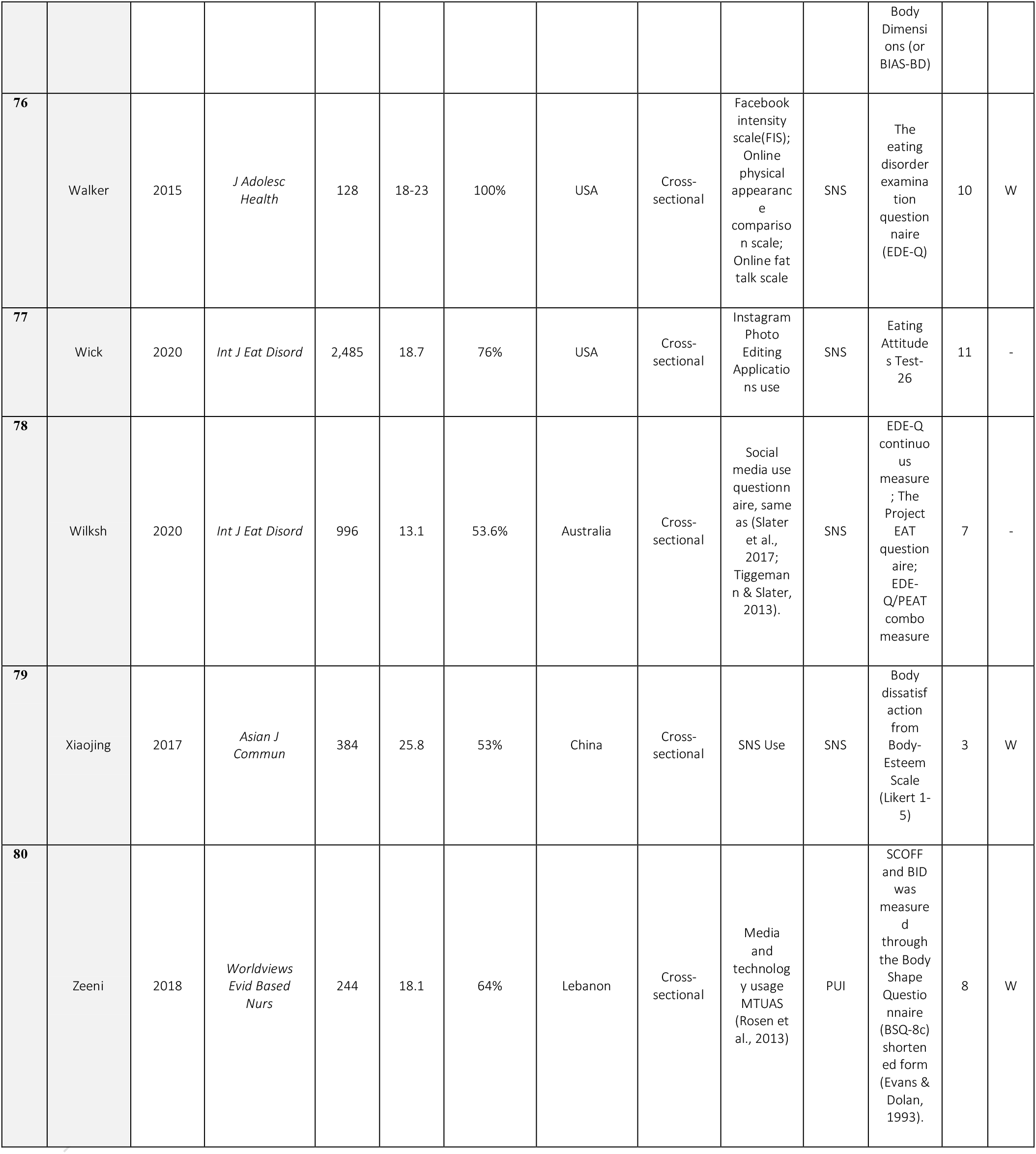
Full list of studies included in the meta-analysis with demographic and problematic internet use characteristics. Full list of studies are presented here in **Table S1** (Ahadzadeh et al., 2017; Almenara et al., 2019; Alpaslan et al., 2015; Aparicio-Martinez et al., 2019; Bair et al., 2012; Bennett et al., 2019; Brown and Tiggemann, 2016; Butkowski et al., 2019; Canan et al., 2014; Cañon Buitrago et al., 2016; Carrotte et al., 2015; Carter et al., 2017; Çelik et al., 2015; Cohen et al., 2017; Cohen and Blaszczynski, 2015; de Vries et al., 2019, 2016; Eckler et al., 2017; Embacher Martin et al., 2018; Fardouly et al., 2015; Fardouly and Vartanian, 2015; Ferguson et al., 2014; Fernández-villa et al., 2015; Griffiths et al., 2018a, 2018b; Hendrickse et al., 2017; Holland and Tiggemann, 2017; Hsieh et al., 2018; Hummel and Smith, 2015; Ivezaj et al., 2017; Kamal and Kamal, 2018; Kelly et al., 2018; Kim and Chock, 2015; Kvardova et al., 2020; Lee et al., 2014; Levinson et al., 2017; Linardon and Messer, 2019; Lonergan et al., 2019; Mabe et al., 2014; Manago et al., 2015; Marco et al., 2018; Martínez-González et al., 2014; McLean et al., 2015; Meier and Gray, 2014; Melioli et al., 2015; Niu et al., 2020; Olenik-Shemesh and Heiman, 2017; Park and Lee, 2017; Pistella et al., 2019; Prichard et al., 2020; Quesnel et al., 2018; Rodgers et al., 2020, 2019, 2013, 2012; Sidani et al., 2016; Simpson and Mazzeo, 2017; Slater et al., 2019, 2017; Smith et al., 2013; Stronge et al., 2015; Sugimoto et al., 2020; Tao, 2013; Tao and Liu, 2009; Terhoeven et al., 2020; Theis et al., 2012; Tiggemann et al., 2020; Tiggemann and Miller, 2010; Tiggemann and Slater, 2017; Tiggemann and Zaccardo, 2015; Tran et al., 2019; Veldhuis et al., 2018; Verbist and Condon, 2019; Wagner et al., 2016; Walker et al., 2015; Wick and Keel, 2020; Wilksch et al., 2020; Xiaojing, 2017; Zeeni et al., 2018). **AN =** Anorexia Nervosa; **BN** = Bulimia Nervosa; **BED** = Binge Eating Disorder; **BSQ =** Body Shape Questionnaire; **CIAS:** Chen Internet Addiction Scale; **DCIA-C:** Diagnostic Criteria for Internet Addiction for College Students; **DSM:** Diagnostic and statistical Manual; **EAT-26 =** Eating Attitudes Test 26 items; **EDE-Q =** Eating Disorder Examination-Questionnaire; **EDI =** Eating Disorder Inventory; **EPHPP** = Effective Public Health Practice Project Quality Assessment Tool for Quantitative Studies; **IAT:** Young’s Internet addiction test; **IADQ:** Young’s Internet Addiction Diagnostic Questionnaire; **Pro-ED** = Promoting Eating Disorder content; **PUI:** Problematic use of the internet; **SATAQ** = Sociocultural Attitudes Towards Appearance Questionnaire-3; **SCOFF =** Sick, Control, One, Fat, Food questionnaire; **SNS** = Social Networking Sites; **VAS**: Visual Analogue scale **VAT:** Video game Addiction Test; **YDQ:** Young’s diagnostic questionnaire; **UWCBs** = Unhealthy weight control behaviors

**TABLE S2.** Ad-hoc quality score system.

| Quality scoring system |  |  |  |
| --- | --- | --- | --- |
|  | Question | Point system | SCORING DETAILS |
| Study design, representativeness and completeness of reported metrics | Q1 - DMG (Age) | reporting age metrics (0-1) | Age mean and standard deviation (or equivalent metrics e.g. median, standard error) need to be reported; otherwise score 0 |
|  | Q2 - DMG (Gen) | reporting gender metrics (0-1) | Distribution between genders need to be reported either by number or percentage or by sample (e.g. 100% males/females) |
|  | Q3 - DMG (Edu) | reporting education metrics (0-1) | Score 0 if no metrics or if sample is unclear (e.g. sampled from X University) but with no metrics; score 1 if specific metrics of education reported or sample group is specified (e.g. sample of College students from X University) |
|  | Q4 - DMG (Eth) | reporting ethnicity metrics (0-1) | Score 1 if distribution between ethnic groups is reported either by number or percentage or by sample (e.g. 100% Caucasian); score 0 if distribution is implied by sample (e.g. students from USA college) but not explicitly stated or if no ethnicity reported |
|  | Q5 - (NOQ-Rep) | representativeness of the exposed cohort (0-2) | Sample is representative of the average in the community, randomized, stratified sampling score 2; volunteer sampling, online survey, selected subgroup, convenience sample, score 1; not defined or not rep sample, score 0 |
| Validity and clarity of reporting of eating disorder and problematic usage of the internet facets | Q6 - PUI (Diagn) | reporting PUI diagnostic metrics (0-2) | Score 2 if diagnostic interview or established PUI clinical sample from records; score 1 if a valid tool of PUI is used; score 0 if unvalidated or only tests PUI by virtue of a single measure of online activity (e.g. n of Facebook friends, time spend online) |
|  | Q7 - PUI (Facet) | reporting PUI facets (0-1) | Score 1 if the facet of PUI (e.g. gaming, social media, or global PUI) are clearly assessed and reported. Score 0 if not or facet unclear |
|  | Q8 - PUI (Out) | reporting PUI outcomes (0-1) | Score 1 if outcome metrics are clearly reported for PUI and non PUI groups or cohort; score 0 if outcomes are not clearly reported. If authors use regression analysis with continuous score, they should also comment on the level above which they consider outcome measure as problematic |
|  | Q9 - ED (Diagn) | reporting ED diagnostic metrics (0-2) | Score 2 if valid tool of eating disorder psychopathology is measured e.g. diagnostic interview (MINI, DSM-4/5, clinical interview); score 1 if valid measure is used; score 0 if ED measure is unclear or only tests limited aspect of ED-symptomatology |
|  | Q10 - ED (Facet) | reporting ED clinical facets (0-1) | Score 1 if the clinical facet of ED (e.g. anorexia nervosa, bulimia, BED, ARFID) are clearly assessed and reported. Score 0 if not or facet unclear |
|  | Q11 - ED (Out) | reporting ED outcomes (0-1) | Score 1 if outcome metrics are clearly reported for ED and non ED groups or cohort; score 0 if outcomes are not clearly reported. If authors use regression analysis with continuous score, they should also comment on the level above which they consider outcome measure as problematic |
| Mental health Confounders | Q12 - Com (MD) | reporting comorbid mood disorders (0-1) | Score one point if common comorbidities have been assessed: Mood disorders (any of depression, bipolar, suicidality, self-injurious behav) |
|  | Q13 - Com (AD) | reporting anxiety disorders (0-1) | Score one point if common comorbidities have been assessed: Any of Anxiety dis (OCD, GAD, Social Phobia, Hypochondriasis, somatization disorder) |
|  | Q14 - (BDD) | reporting BDD (0-1) | Score one point if common comorbidities have been assessed: Body dysmorphic disorder |
|  | Q15 - (EA) | reporting Exercise addiction (0-1) | Score one point if common comorbidities have been assessed: Exercise Addiction |
|  | Q16 - (SUD) | reporting substance misuse (0-1) | Score one point if common comorbidities have been assessed: Substance misuse (any) |
|  | Q17 - (ICD) | reporting ICDs (0-1) | Score one point if common comorbidities have been assessed: Impulse control disorders (kleptomania, gambling, trichotillomania, pyromania, IED etc.) |
| Neurobiological determinants | Q18 - (Beh) | reporting neurobiological determinants (0-1) | Score one point if the papers reports any neurobiological measure of behaviors (e.g. impulsivity, compulsivity, obsessionality, intolerance to uncertainty, sensation seeking or other) linked both to PUI and ED |
|  | Q19 - (CSF) | reporting neurobiological determinants (0-1) | Score one point if the papers reports any neurobiological measure of cognition of brain structure/function linked both to PUI and ED |

**TABLE S3.** Heterogeneity and model estimate measures for different domains of eating disorder. **tau^2**: estimated amount of total heterogeneity; **I^2**: (total heterogeneity / total variability); **H^2**: (total variability / sampling variability); **Q-test**: Test for Heterogeneity; meta-analysis was done using random-effects model using REML. **REML**: Restricted maximum-likelihood estimator

| Domain | N studies | $\tau^2$ (se) | $I^2$ | $H^2$ | Q-test (p value) † | Model Estimate (se) | sig. |
| --- | --- | --- | --- | --- | --- | --- | --- |
| At-risk eating disorders | 16 | 0.0216<br>(SE = 0.0086) | 93.97% | 16.59 | < .0001 | 0.21 (0.038) | *** |
| Body dissatisfaction (all studies) | 24 | 0.0075<br>(SE = 0.0030) | 82.14% | 5.60 | < .0001 | 0.16 (0.02) | *** |
| Body dissatisfaction (males only) | 4 | 0.0054<br>(SE = 0.0073) | 66.36% | 2.97 | 0.025 | 0.12 (0.047) | ** |
| Drive for thinness | 9 | 0.0077<br>(SE = 0.0062) | 68.62% | 3.19 | 0.012 | 0.16 (0.037) | *** |
| Dietary restraint | 6 | 0.0040<br>(SE = 0.0044) | 54.92% | 2.22 | 0.0375 | 0.18 (0.033) | *** |

**TABLE S4.** Heterogeneity and model estimate measures for different domains without exclusion of influential cases. **tau^2**: estimated amount of total heterogeneity; **I^2**: (total heterogeneity / total variability); **H^2**: (total variability / sampling variability); **Q-test**: Test for Heterogeneity; meta-analysis was done using random-effects model using REML. **REML**: Restricted maximum-likelihood estimator

| Domain | N studies | $\tau^2$ (se) | $I^2$ | $H^2$ | Q-test (p value) † | Model Estimate (se) | sig. |
| --- | --- | --- | --- | --- | --- | --- | --- |
| At-risk eating disorders | 17 | 0.0558<br>(SE = 0.0202) | 97.50% | 40.04 | < .0001 | 0.26 (0.05) | *** |
| Body dissatisfaction (mixed) | 24 | 0.0075<br>(SE = 0.0030) | 82.14% | 5.60 | < .0001 | 0.16 (0.02) | *** |
| Body dissatisfaction (males only) | 4 | 0.0054<br>(SE = 0.0073) | 66.36% | 2.97 | 0.025 | 0.12 (0.047) | ** |
| Drive for thinness | 10 | 0.0275<br>(SE = 0.0153) | 89.56% | 9.58 | < .0001 | 0.20 (0.057) | *** |
| Dietary restraint | 6 | 0.0040<br>(SE = 0.0044) | 54.92% | 2.22 | 0.0375 | 0.18 (0.033) | *** |

**Table S4.**
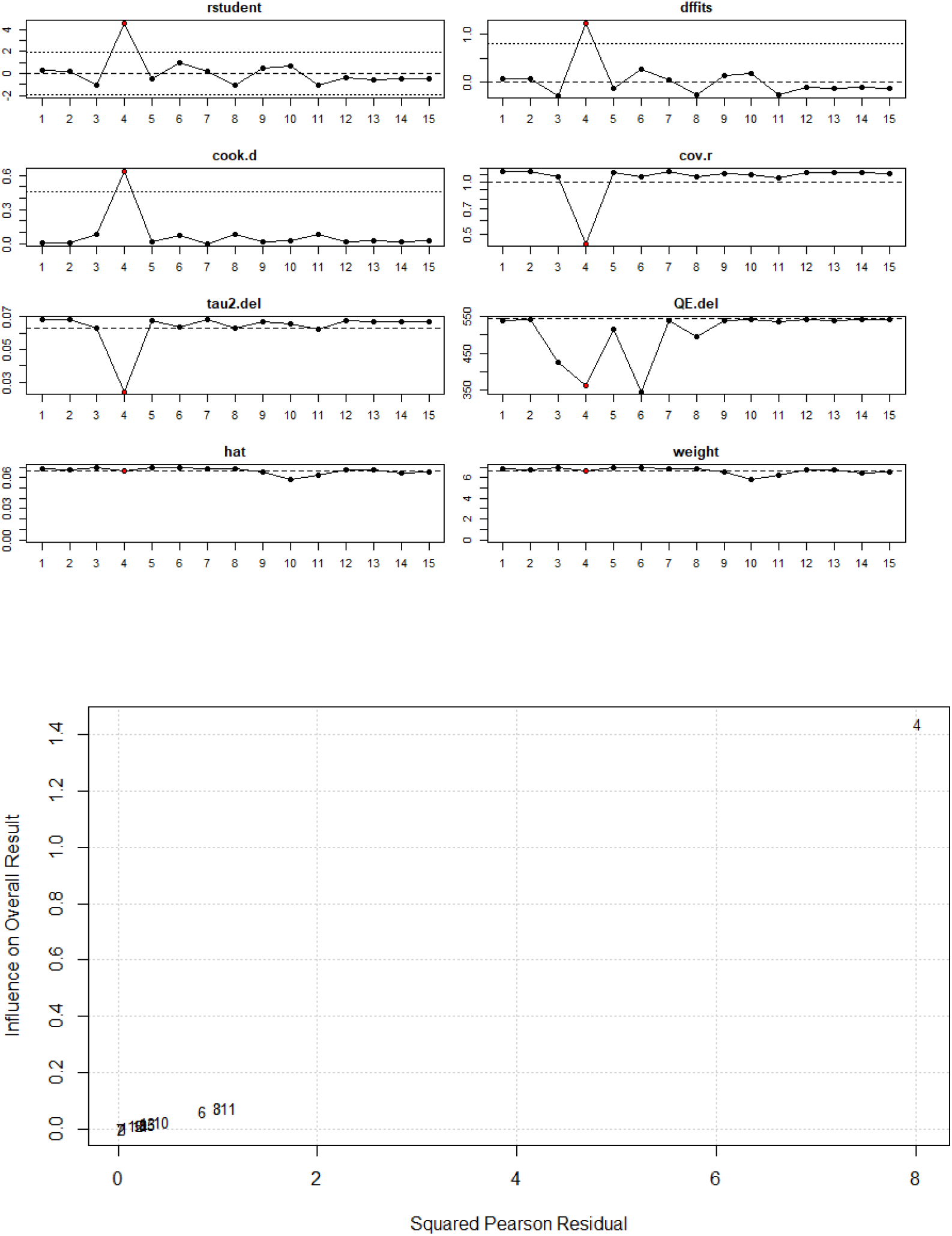
Influence plots describing the influential effects of studies (pre-exclusion) At-risk eating disorders: Outlier and influential studies diagnostics. Study ID = 4 Celik et al. (Çelik et al., 2015)

**Table S5.**
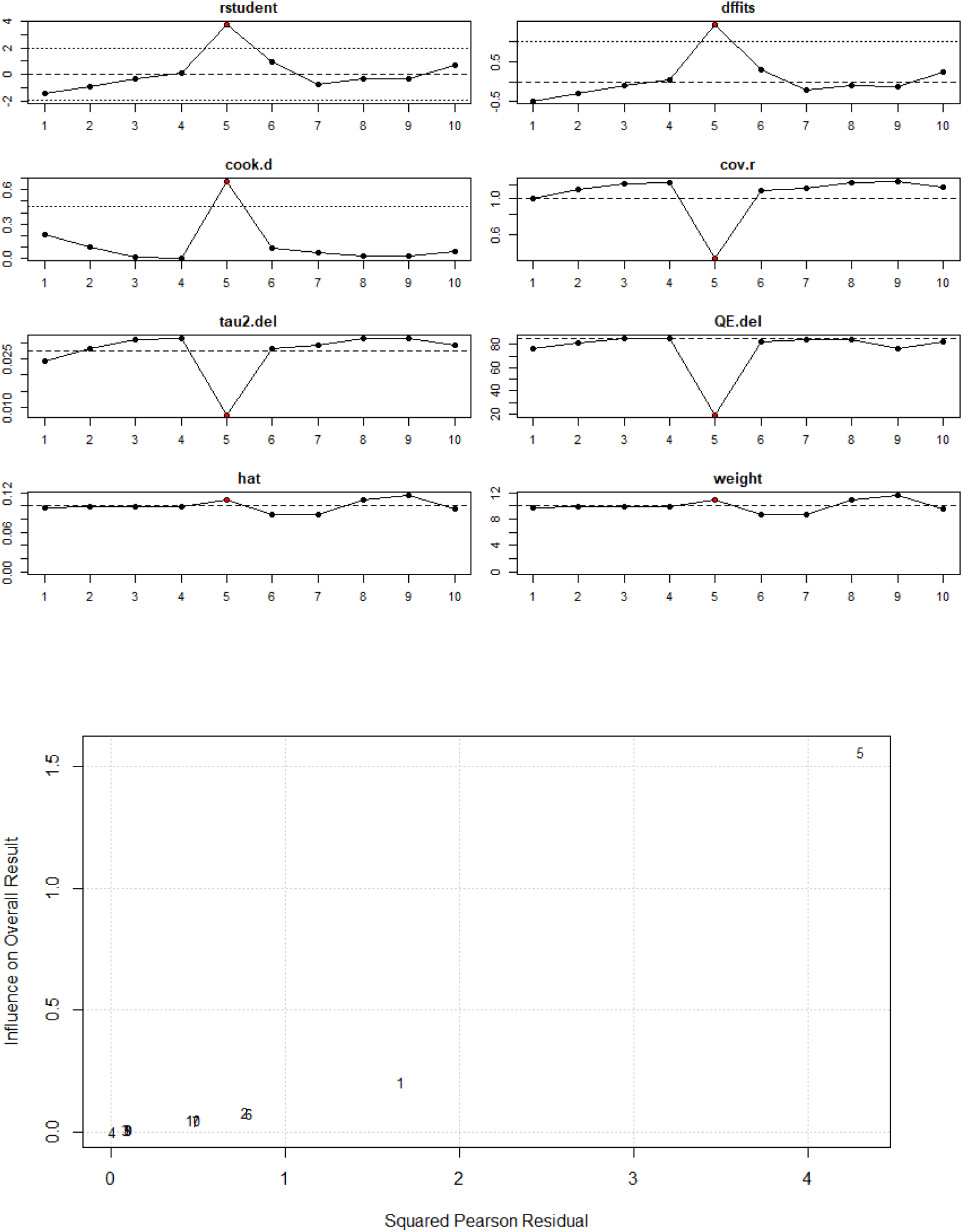
Influence plots describing the influential effects of studies (pre-exclusion) **Drive for Thinness:** Outlier and influential studies diagnostics. Study ID = 5 Kvardova et al. (Kvardova et al., 2020).

## ∫ – S1 Metrics of Problematic usage of the internet

The Internet Addiction Test (IAT) (Young, 1998) provides a measure of maladaptive internet use comprising 20 questions examining multiple facets of PUI, whereas a more modern 18-item instrument, the Problematic Internet Use Questionnaire (PIUQ), has been demonstrated to have excellent psychometric properties and factorial structure (Demetrovics et al., 2008). The Compulsive Internet Use Scale is a short, psychometrically sound questionnaire of problematic internet use comprising 14-Likert-type-items (Meerkerk et al., 2009). The Chinese Internet Addiction Scale is a widely used instrument in Internet Addiction research with good test-retest reliability and internal consistency (Chen et al., 2003). Despite the existence of standardized instruments with good psychometric properties, many studies utilized ad-hoc and application or site specific instruments to ascertain the degree or severity of the online usage behavior. For example, many studies used metrics ascertaining duration of use (time spent on Facebook, Instagram, Snapchat, Tumblr, appearance-focused internet gaming) or extend of specific behavior (number of friends, number of posts, number of accounts, visits per day), or ascertaining behavior that is deemed maladaptive (e.g. seeking negative social evaluations and/or engagement in social comparisons)(Smith et al., 2013). Other SNS based measures included the Facebook Questionnaire (FBQ;(Meier and Gray, 2014), Social Networks Addiction Questionnaire (SNSA) (P. et al., 2001), the Multidimensional Scale of Facebook Use (MSFU; (Frison and Eggermont, 2016)), Social network usage and appearance comparisons (Fardouly and Vartanian, 2015).

## ∫ – S2 Metrics of eating disorders

### S2.1 Eating Attitudes Test

The Eating Attitudes Test (EAT-40 & EAT-26) (Garner et al., 1982) is a reliable, valid and economical instrument useful as an objective measure of the symptoms of anorexia nervosa. The global score cut-off of 20 or above has been used to described disordered eating; the more stringent cut-off of 30 and above has been used as well, as it correlates highly with clinical eating disorder diagnoses.

### S2.2 Eating Disorder Inventory

The Eating Disorder Inventory (EDI) (Garner et al., 1983) is a self-report questionnaire used to assess the presence of eating disorders. The latest version (EDI-3) comprises nine psychological subscales for eating disorders and yields six composite scores. It includes subscales for Drive for Thinness (EDI-DT), Bulimia (EDI-B) and Body Dissatisfaction (EDI-BD).

### S2.3 SCOFF Questionnaire

The SCOFF Eating Disorder Questionnaire (Luck et al., 2002) is a validated screening tool for detection of eating disorders, specifically either anorexia nervosa or bulimia nervosa. A score of 2 and above indicates high likelihood of anorexia nervosa or bulimia nervosa.

### S2.4 Eating Disorder Examination Questionnaire

The Eating Disorder Examination Questionnaire EDE-Q (Fairburn and Beglin, 1994) is widely used measure of eating disorder psychopathology; it assesses the range and severity of eating disorder features. The EDE-Q has four subscales: a) Restraint b) Eating Concern, c) Weight Concern, d) Shape Concern and a Global Score.

### S2.5 Body dissatisfaction

Body Image Avoidance Questionnaire (BIAQ) (Rosen et al., 1991) is a s a self-report measure of behavioral avoidance of experiences that could increase body image-related distress or dissatisfaction. The Body Shape Questionnaire (BSQ) is a self-report questionnaire that was developed to measure concerns about body shape (Cooper et al., 1987). The body dissatisfaction subscale of the Body Attitudes Test (BAT; Probst et al. 1995) is a measure body dissatisfaction (Probst et al., 1995). The Body Image States Scale (BISS) is a six-item scale of body image evaluation (Cash et al., 2002). The Contour Drawing Rating Scale (CDRS) difference score is aimed to ascertain the difference between perceived and desired body shape (Thompson and Gray, 1995). Other studies used manifest scales specifically made for the purposes of their study e.g. the body dissatisfaction scale for the New Zealand Attitudes and Values Study (NZAVS) questionnaire battery (Stronge et al., 2015).

### S2.6 Dietary restraint

The Dutch Eating Behavior Questionnaire (DEBQ) (Strien et al., 1986) has high internal consistency and factorial validity as well as external validity and contains a 10-item restraint subscale (DEBQ-R). Studies have used DEBQ-R or EDE-Q restraint subscale.

## R code used in the analysis

\# Use the following the command to load the data. Only sample of code is presented here, repeated sequences of code are omitted.

\# For the analyses described in Quintana 2015 (Quintana, 2015), the data is included with the metafor package. Use the following the command to load the data. You are creating a new object called “dat”

df <- read.csv(“∼/PhD CAPHRI/XYZ/meta_data.csv”, head = TRUE, stringsAsFactors = FALSE); df <- data.table(df)

df <- df %>% filter(Analysis.method = = “Pearson”); str(df)

#df <- mutate(df, study_id = 1:92) # This adds a study id column

df_r_ared <- df %>% dplyr::select(id, author,Year, n,PUI_facet, r_ared) %>%

filter(complete.cases(.))# %>% df_r_ared$r_ared <- as.numeric(df_r_ared$r_ared)

df_r_bd <- df %>% dplyr::select(id, author,Year, n,PUI_facet, gender, r_bd) %>% filter(complete.cases(.))# %>%

df_r_bd$r_bd <- as.numeric(df_r_bd$r_bd)

df_r_dt <- df %>% dplyr::select(id, author,Year, n,PUI_facet, r_dt) %>%

filter(complete.cases(.))# %>% df_r_dt$r_dt <- as.numeric(df_r_dt$r_dt)

df_r_bn <- df %>% dplyr::select(id, author,Year, n,PUI_facet, r_bn) %>%

filter(complete.cases(.))# %>% df_r_bn$r_bn <- as.numeric(df_r_bn$r_bn)

df_r_re <- df %>% dplyr::select(id, author,Year, n,PUI_facet, r_re) %>%

filter(complete.cases(.))# %>% df_r_re$r_re <- as.numeric(df_r_re$r_re)

\# The first step is to transform r to Z and calculate the corresponding sample variances.

#to exclude influential run this; if not ru witout: df_r_ared <- df_r_ared %>% filter(id ! = “8”); dat <-escalc(measure = “ZCOR”, ri = r_ared, ni = n, data = df_r_ared, slab = paste(author, Year, sep = “, “)); res <- rma(yi, vi, data = dat); res; predict(res, digits = 3, transf = transf.ztor); confint(res)

b_res <- rma(yi, vi, data = dat, slab = id) # New meta-analysis with study ID identifier

\# The next command will plot a Baujat plot.; baujat(b_res)

\# Studies that fall to the top right quadrant of the Baujat plot contribute most to both these factors. Looking at the Molloy et al., 2014 data set reveals that 3 studies that contribute to both of these factors. A closer look the characteristics of these studies may reveal moderating variables that may contribute to heterogeneity

\# A set of diagnostics are also available to identify potential outliers and influential cases.

inf <- influence(res); print(inf); plot(inf) # The plot visualizes the printed dataset. As there are no studies are marked with an asterisk in the printed dataset, none of the studies fulfilled the criteria as an influential study. # Now we visualize the meta-analysis with a forest plot.

forest(res, xlim = c(−1.6,1.6), atransf = transf.ztor, at = transf.rtoz(c(-.4,-.2,0,.2,.4,.6)), digits = c(2,1), cex =.8)

text(−1.6, 18.5, “Author(s), Year”, pos = 4, cex =.9); text(−0.3, 18.5, “At-risk-eating-disorders”, pos = 4, cex =.9); text(1.6, 18.5, “Effect size [95% CI]”, pos = 2, cex =.9)#repeat code for different domains

#moderator analysis; df <- df %>% select(ID, paper, Age:Geographical, Co.morbidities:control.N) # This brings the study id column to the front

df$Age <- as.factor(df$Age); df$Co.morbidities

<- as.factor(df$Co.morbidities)

dfm_Stroop <- df %>% filter(Task = = “Stroop”) %>%

dplyr::select(ID:control.N) %>% filter(complete.cases(.))

res.modage <- rma(yi, vi, mods = ∼ Age, data = dfm_Stroop);res.modage

res.modgender <- rma(yi, vi, mods = ∼ Gender, data = dfm_Stroop);res.modgender

res.modgeo <- rma(yi, vi, mods = ∼ Geographical, data = dfm_Stroop);res.modgeo

res.modmorb <- rma(yi, vi, mods = ∼ Co.morbidities, data = dfm_Stroop);res.modmorb (…) #repeat by domain

